# Quantitative Microbial Risk Assessment of Extended-Spectrum β-Lactamase-Producing *Escherichia coli* Transfer from Broiler Litter to Fresh Lettuce Consumption

**DOI:** 10.1101/2025.06.28.25330457

**Authors:** Nunzio Sarnino, Subhasish Basak, Lucie Collineau, Roswitha Merle

**Affiliations:** Freie Universität Berlin, Institute of Veterinary Epidemiology and Biostatistics, Veterinary Centre for Resistance Research Berlin, Germany; University of Lyon - French Agency for Food, Environmental and Occupational Health and Safety (ANSES), Epidemiology and Surveillance Support Unit,Lyon, France

## Abstract

**Background:** Extended-spectrum β-lactamase-producing *Escherichia coli* (ESBL *E. coli*) from broiler chicken production pose potential public health risks via multiple environmental and foodborne pathways. We developed a modular quantitative microbial risk assessment (QMRA) model linking four components, namely farm, soil, river, and lettuce consumption, to predict human environmental exposure to ESBL *E. coli* originating from broiler flocks.

**Methods:** A stochastic farm module simulated broiler colonization over a 36-day cycle and generated end-cycle litter loads. Field modules represented first-order decay, attachment/partitioning, and runoff to rivers; irrigation transfer yielded lettuce contamination for a 100 g serving. Disability-Adjusted Life Year (DALY) estimation characterized health risk and global sensitivity analyses identified main drivers.

**Results:** The farm model produced mean end-cycle litter of 1.6 × 10^4^ (SD 16.1, UI 1.60–1.61 × 10^4^) CFU/g and near-complete flock colonization within one week. Soil surface loads declined from 3.2 × 10^7^ (SD 3.2 × 10^4^, UI 3.2–3.2 × 10^7^) CFU/m^2^ to 8.6 × 10^5^ (SD 8.8 × 10^2^, UI 8.6–8.6 × 10^5^) CFU/m^2^ by day 100. Runoff yielded river concentrations of 6.0 × 10^-2^ (SD 3.5 × 10^-3^, UI 5.5–6.6 × 10^-2^) CFU/mL after ten days. Lettuce contamination was highest at short intervals after land application 1.7 (SD 8.5 × 10^-2^, UI 1.5–1.8) CFU/100 g at one day interval, dropping to 0.85 (SD 5.0 × 10^-2^, UI 0.76–0.95) CFU/100 g at day 20 interval; simple household washing cut exposure by ∼90 %. Global sensitivity analysis identified soil-water partitioning and decay rates as most important parameters of exposure variability. DALYs lost ranged from 10^-8^ to 10^-10^ per serving.

**Conclusions:** In our scenarios, predicted health burdens varied with ESBL *E. coli* concentration in irrigation water and intervals between litter application and lettuce planting. While environmental decay and simple measures such as household washing substantially reduce exposure, residual contamination persists. Future studies should evaluate the effectiveness of manure treatments and irrigation-water quality interventions on reducing environmental loads and human risk.

## Introduction

Antimicrobial resistance (AMR) is steadily reducing the effectiveness of antibiotics in human and veterinary medicine. In broiler farming, antimicrobial use may create strong selective pressure and accelerate the emergence and spread of resistant bacteria (Chantziaras et al. 2014). A prime example is extended-spectrum β-lactamase (ESBL)-producing *Escherichia coli (E. coli)*, which can break down β-lactam antibiotics. These organisms pose a serious public health threat and could increase healthcare costs (Leistner et al. 2014; MacKinnon et al. 2020; Apostolakos et al. 2019)

Resistant strains may enter broiler houses in multiple ways. Day-old chicks may arrive already colonized (Moreno et al. 2019), or resistance may pass vertically from breeder flocks (Dierikx et al. 2013; Agersø et al. 2014; Nilsson et al. 2014). Environmental contamination and biosecurity lapses offer additional routes (Mallioris et al. 2023; Robé et al. 2024). Although the prevalence of resistance at chick placement is generally low (Oikarainen et al. 2019), within-flock spread frequently leads to near complete colonization within a few days (Robé et al. 2019).

Once broiler houses are emptied, litter used as fertilizer potentially serves as a reservoir of ESBL *E. coli*. These bacteria can survive in soil for extended periods (van Overbeek et al. 2021), enter surface and groundwater through runoff (Sowah et al. 2020), and contaminate produce (Jensen et al. 2013; Mootian et al. 2009). Humans may then be exposed via contaminated food, recreational water contact, or other environmental pathways (O’Flaherty et al. 2018; O’Flaherty et al. 2019a; O’Flaherty et al. 2019b; Chapman et al. 2018). Environmental factors, notably rainfall events, can significantly influence bacterial concentrations in rivers, often leading to transient peaks (Alegbeleye and Sant’Ana 2020; Jacobs et al. 2019). Furthermore, the proximity to animal production sites seems to be an additional risk factor (Hubbard et al. 2020; Chapman et al. 2018).

Litter management may reduce ESBL *E. coli* loads. Thermophilic composting inactivates pathogens and lowers bacterial counts (Thomas et al. 2024; Okada et al. 2024). Respecting pre-harvest intervals limits the environmental spread (Subirats et al. 2021). Advanced composting and short-term litter storage further reduce resistant bacterial populations (Atanasova et al. 2025; Siller et al. 2020; Howard et al. 2016; Suzuki et al. 2024). However, bacterial survival depends on environmental conditions, underscoring the need for site-specific risk assessments and tailored interventions (Çekiç et al. 2017; van Overbeek et al. 2021; Sarnino et al. 2025).

Quantitative microbial risk assessment (QMRA) frameworks offer a systematic approach to characterize the propagation of resistant pathogens and to evaluate the potential effect of control measures (Collineau et al. 2020; Rawson et al. 2019; Fastl et al. 2023).

Within the ENVIRE project (https://www.envire-project.de/), we developed an integrated QMRA model to simulate how ESBL *E. coli* move from broiler farms under European conditions into the environment and reach humans. Our objectives were to:

1. Predict ESBL *E. coli* loads in raw broiler litter at the end of a conventional production cycle,
2. Characterize transmission pathways from litter to soil, water, and crops,
3. Identify the dominant drivers of exposure at consumption and how the time between litter application and planting shapes contamination at harvest,
4. Estimate human exposure and Disability-Adjusted Life Year (DALYs) lost via lettuce consumption.

Other potential routes (for example direct crop contamination from soil) were not included because our previous work (Sarnino et al. 2025) found that crop contamination primarily occurs via irrigation water.

This study is, to our knowledge, the first modular Quantitative Microbial Risk Assessment (QMRA) to connect broiler farm, amended soil, receiving water, lettuce consumption for ESBL *E. coli*, using one internally consistent set of assumptions across modules. We (i) link flock colonization dynamics to end-cycle litter loads, (ii) propagate soil partitioning and hydrological wash-off to river concentrations, (iii) map irrigation transfer to edible portions, and (iv) quantify health burden (DALY). This integrated framing extends prior single-pathway models and generic *E. coli* assessments by targeting ESBL strains and explicitly testing time-since-application scenarios relevant to produce safety.

## Materials and Methods

### Model Description

We built a modular QMRA model to estimate human exposure to ESBL *E. coli* from broiler production. The model links four modules - farm, soil, river and lettuce consumption - to trace contamination from its source all the way to human exposure (Figure 1).

**Figure 1.**
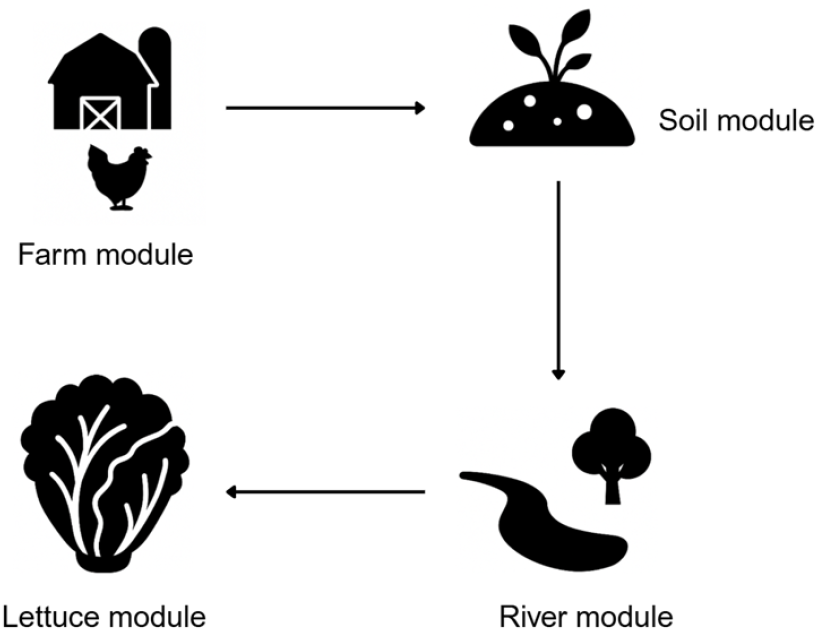
Diagram of the QMRA model linking the farm, soil, river, and lettuce modules.

In the farm module, we simulated a 36-day broiler cycle with a stochastic susceptible-infectious compartmental model. Broilers enter as susceptible (*S*) or colonized (*C*) at a set prevalence (*p*_init_) (Table 1). Gut bacterial growth (*r*) follows a logistic curve; infection may spread via ingestion (*ρ*_ingest_) of contaminated feces 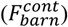. Each day, the model updates broiler age, infection duration, fecal output, and bacterial shedding. ESBL *E. coli* levels accumulate in the barn litter and then decline by natural decay.

**Table 1.** Key input variables for the farm module.

| Variable | Description | Source | Unit | Value |
| --- | --- | --- | --- | --- |
| $\alpha$ | Proportion of ingested water lost by metabolism | Becker et al. (2022) | proportion | 0.7 |
| $\epsilon$ | Excretion rate of ESBL *E. coli* | Becker et al. (2022) | % | 0.3 |
| $D_{\max}$ | Total duration of the fattening period | Assumed | day | 36 |
| $r_{\min}$ | Minimum growth rate for ESBL *E. coli* in the broiler's intestine | Becker et al. (2022) | log10 CFU | 0 |
| $r_{\max}$ | Maximum growth rate for ESBL *E. coli* in the broiler's intestine | Becker et al. (2022) | log10 CFU | 5 |
| $K$ | Maximum amount of bacteria in the substrate (the intestinal content in this case) | Robé et al. (2019) | CFU/g | $10^6$ |
| $\phi$ | Environmental decay rate of ESBL *E. coli* in broiler litter | Becker et al. (2022) | % | 0.5 |
| $\rho_{\text{ingest}}$ | Proportion of ingested contaminated feces along with feed | Becker et al. (2022) | % | 0.014 |
| $\beta$ | Transmission rate | Dame-Korevaar et al. (2020) | CFU/day | 0.31 |
| $w_{\min}(d)$ | Minimum water consumption on day $d$ | Avian advice 11(2), University of Arkansas. (2009) | g | 21.16–10.35 |
| $w_{\max}(d)$ | Maximum water consumption on day $d$ | Avian advice 11(2), University of Arkansas. (2009) | g | 42.66–31.18 |
| $g(d)$ | Broiler's average daily gain in weight on day $d$ | Ross308 performance objectives. (2022) | g | 21–100 |
| $I(d)$ | Broilers average feed intake on day $d$ | Ross308 performance objectives. (2022) | g | 20–189 |
| $A$ | Size of the farm | Assumed | m<sup>2</sup> | 100 |
| $\rho_{\text{farm}}$ | Stocking density | Directive 2007/43/EC — minimum rules for the protection of chickens kept for meat production | kg/m<sup>2</sup> | 39 |
| $w_{\text{tar}}$ | Weight of the broilers at the end of the fattening period (36-th day) | Becker et al. (2022) | kg | 2.332 |
| $c_{\text{init}}$ | Total CFU of ESBL *E. coli* in colonized one day old chicks | Becker et al. (2022) | CFU | 100 |
| $p_{\text{init}}$ | Initial prevalence of colonized broilers | Expert opinion, Robé | Fraction | 0.01 |
| $L$ | Litter quantity per square meter | Becker et al. (2022) | g/m<sup>2</sup> | 1000 |

Throughout this article, we use the term “litter” to refer to the mixture of broiler manure, bedding, feathers, and other materials commonly removed at the end of a production cycle, while “clean litter” is the fresh bedding material added at the start of a production cycle.

The soil module applies broiler litter to an agricultural field - assuming zero delay between chicken harvest and litter spread - and models ESBL *E. coli* decay using an exponential rate fitted to experimental data from Sharma Manan et al. (2019). While poultry litter is often treated, this is not a universal or legally mandated practice across the EU. There are no specific regulations requiring all poultry litter to be treated or specifying an interval between removal from the barn and field application. Therefore, in our model, we consider a worst-case scenario in which broiler litter is applied to fields immediately after removal from poultry houses.

The river module estimates daily bacterial runoff from the field to the river using a simplified SWAT (soil and water assessment tool)-inspired approach (Douglas-Mankin et al. 2010). We considered water temperature, depth, solar radiation, and salinity, then applied wash-off fractions and in-water decay to predict river concentrations.

The lettuce module simulates multiple planting events over a growing season. On irrigation days, bacteria adhere to leaves in proportion to river concentration and retention volume. From irrigation to harvest, loads decrease according to a biphasic decay model, and post-harvest washing further reduces bacterial counts. Exposure is reported as colony-forming units per 100 g serving. In this module, we assume that the lettuce is consumed on harvest day.

Across all modules, we employed a stochastic approach; each module ran 1000 simulations to reflect natural variability in environmental conditions and individual behaviour. For each result, we defined the standard deviation (SD) as descriptive measure of dispersion around the mean and the 95% uncertainty interval (UI) as the 2.5th and 97.5th percentiles of the resulting empirical distribution.

### Model credibility

We harmonized units and variable definitions across module, constrained inputs to ranges reported in peer-reviewed studies or agronomic practice, and ran all analyses from scripted, version-controlled code. To support external validity, we planned piecewise checks against independent evidence, comparing our results with experimental studies and other QMRA.

### Farm Module

The farm module simulates the transmission dynamics of ESBL *E. coli* between two primary reservoirs: the broilers’ gastrointestinal tract and the litter within the barn environment. A single simulation of this stochastic module corresponds to a complete broiler production cycle of *D*_max_ days of a single flock of broilers. The module operates within a discrete-time framework with a daily time step. For each day *d* = 1, 2, …, *D*_max_ of the production cycle, a series of barn-level events, each representing distinct mechanisms in the transmission dynamics of ESBL *E. coli* within the barn environment, is executed once per day. These events form the core components of the farm module and are detailed following the model initialization.

### Module initialization

The farm module is initialized using the total number of broilers in a flock computed as,

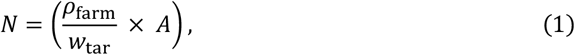

where *ρ*_farm_ is the farm stocking density (kg/m^2^), *w*_tar_ is the target weight (in kg) at day *D*_max_, and *A* is the farm area (m^2^). Further let *N*_col_(*d*), for *d* = 1, 2, …, *D*_max_, denote the total number of colonized broilers in the flock on day *d*, with an initial proportion *p*_init_ used to set *N*_col_(1). The flock is partitioned into two compartments: susceptible (*S*) and colonized (*C*), with no recovery assumed during the production cycle; their respective initial sizes are (*N* − *N*_col_(1)) and *N*_col_(1).

For *i* ∈ *C* ∪ *S* and *d* = 1, 2, …, *D*_max_, let 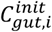 (*d*) denote the initial total number of CFUs of ESBL *E. coli* present in the *i*-th broiler’s gut on day *d*. 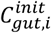 (1) is set to 0 CFU for every *i* ∈ *S* and 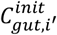(1) is set to *c*_init_ CFUs, for every *i*^*′*^ ∈ *C*. Moreover, on day 1, the barn litter is assumed to be completely free of ESBL *E. coli*, with no CFUs present. Consequently, the value of both 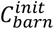 (1) and 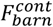 (1), representing respectively the total initial CFU count and amount of contaminated feces in the litter on day 1, are set to zero.

### Feces excretion

The first barn-level event implemented by the farm module is the production of feces by the broilers. On day *d*, for *d* = 1, 2, …, *D*_max_, the *i*-th broiler’s feces output denoted by *F*_*i*_(*d*), for *i* ∈ *C* ∪ *S*, is calculated as,

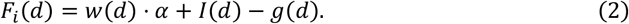

Here *w*(*d*) is a uniformly distributed random variable with limits *w*_min_(*d*) and *w*_max_(*d*), representing respectively the day-specific bounds on water consumption of broilers. *α* is a fractional reduction factor for water. *I*(*d*) and *g*(*d*) respectively denote the average feed intake (kg/day) and daily weight gain (kg/day) of a broiler Ross 308. After the excretion of day, the total amount of contaminated feces in the barn environment till the beginning of day (*d* + 1) is given by,

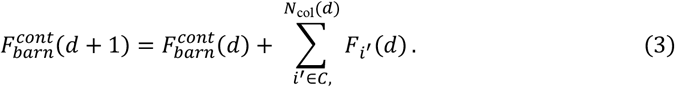

### ESBL *E. coli* excretion

The event ESBL *E. coli* excretion models the excretion of ESBL *E. coli* in the barn environment along with the feces by the colonized broilers. The total number of CFUs of ESBL *E. coli* remaining in the *i*^*′*^-th colonized broiler’s gut, for *i*^*′*^ ∈ *C*, on day *d* after excretion, is given by,

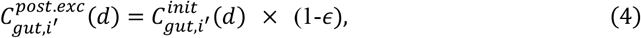

where *ϵ* is the excretion rate. The total cumulated load of ESBL *E. coli* (in CFUs) in the litter after excretion on day *d*, for *d* = 1, 2, …, *D*_max_, is obtained as,

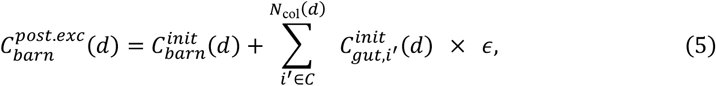

### ESBL *E. coli* growth inside broilers’ guts

Within colonized broilers, bacterial growth event in the gut is modeled with a logistic growth function (see, e.g., Tsoularis and Wallace 2002). With *K* being the carrying capacity and the growth rate 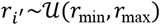 distributed uniformly, the number of CFUs of ESBL *E. coli* in the gut of *i*^*′*^-th colonized broiler, for *i*^*′*^ ∈ *C*, on day *d* after the growth event is given by:

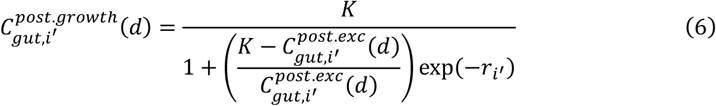

This formulation captures saturation effects and within-flock variability through random draws for 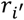.

### Feces ingestion

The next barn-level event is the ingestion of feces where each broiler is assumed to accidentally ingest a certain proportion *ρ*_ingest_ of contaminated fecal material from the barn environment (see, e.g., Becker et al., 2022), along with their daily feed. This triggers the colonization of susceptible, non-colonized broilers, while the already colonized ones remain persistently colonized. The total amount of contaminated feces (in g) accidentally ingested by the *i*-th broiler, for *i* ∈ *C* ∪ *S*, on day *d* is given by,

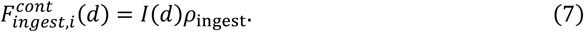

Let *N*_new_(*d*) denote the expected number of newly colonized broilers on day *d* through accidental ingestion of contaminated feces. At this stage, the farm module converts the same number of susceptible broilers to the colonized state by simple random sampling without replacement. Simultaneously at this moment, the flock is re-partitioned into three mutually exclusive and exhaustive compartments, namely, susceptible (*S*), newly colonized (*C*^*^) on day *d*, and previously colonized (*C*), with respective sizes (*N* − *N*_col_(*d*) − *N*_new_(*d*)), *N*_new_(*d*) and *N*_col_(*d*). The quantity of ESBL *E. coli* ingested by already-colonized broilers is considered negligible compared with their gut load and is therefore neglected. The gut concentrations for broilers belonging to all the three compartments are updated as,

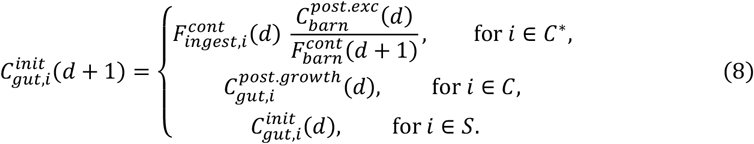

Simultaneously the ESBL *E. coli* load in the barn environment is also updated as,

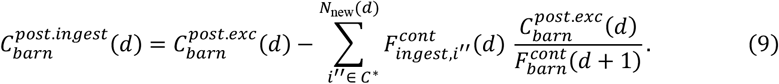

However, the quantity of contaminated feces ingested from the barn environment by all the broilers is considered negligible compared to 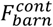 (*d* + 1), and hence it is kept unchanged.

### Transmission

The transmission event is used to estimate *N*_new_(*d*). The framework proposed by Dame-Korevaar (see, e.g., Dame-Korevaar et al. 2020a) is adapted as a conceptual basis but extended to honour flock heterogeneity and mass balance constraints. The module is based on the following assumptions, (1) broilers acquire ESBL *E. coli* only by accidentally ingesting contaminated litter and broiler-to-broiler transmission through direct contact is ignored; (2) the simulation operates on a fixed time-step of 1 day, so each broiler experiences at most one exposure decision per day. The force of infection *λ*(*d*) relates to the environmental concentration of ESBL *E. coli* to *N*_new_(*d*), and is derived as,

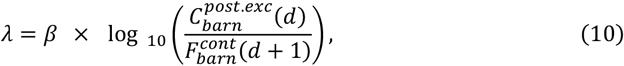

where *β* is the transmission coefficient. The expected number of new colonization during the interval on day *d* is obtained as,

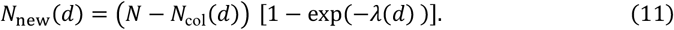

On day (*d* + 1), the two compartments *C*^*^ and *C* are merged as *C* and the corresponding size is updated as *N*_col_(*d* + 1) = *N*_col_(*d*) + *N*_new_(*d*). This ensures that the newly colonized broilers contribute to excretion of contaminated feces and experience growth of ESBL *E*.*coli* inside their guts.

### Environmental decay

The final barn-level event is the environmental decay which models the reduction in ESBL *E. coli* population in the barn environment due to absence of suitable growth conditions and thus preventing indefinite bacterial build-up. For a decay rate *ϕ* the initial barn load for the next day after environmental decay, is obtained as,

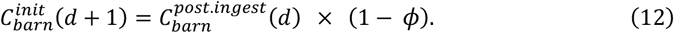

### Final concentration calculation

At the end of a single simulation of the farm module on day *D*_max_, the final ESBL *E. coli* concentration in litter (CFU/g) denoted by *C*_final_, is obtained by dividing the total environmental load by the sum of accumulated feces and the mass of fresh bedding *L*,

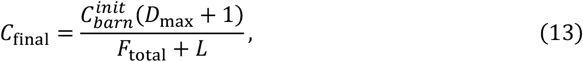

where 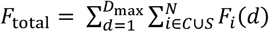 is the aggregated feces mass by day *D*_max_.

### Soil Module

All parameter values and distributional assumptions for the soil and river modules can be found in Table 2.

**Table 2.** Key input variables for the soil, river, exposure and lettuce-exposure modules.

| Variable | Description | Source | Unit | Value |
| --- | --- | --- | --- | --- |
| $k$ | First-order die-off rate of ESBL-E. coli in litter-amended top-soil | Sharma Manan (2019) | /Day | 0.0362 |
| $R_{app}$ | Broiler litter applied to field surface | Assumed | Kg/m<sup>2</sup> | 2 |
| $K_d$ | Bacteria partition coefficient | Sowah (2020) | Fraction | 0.95 |
| $W_i$ | Mobile-phase cells washed off per run-off event | Sowah (2020) | Fraction | 0.50 |
| $T$ | River-water temperature | O’Flaherty 2019a | °C | 21–28 |
| $EC$ | Salinity | O’Flaherty 2019a | % | 0.035–0.075 |
| $IA$ | Global solar irradiance | O’Flaherty 2019a | Ly/h | 17.3–25.4 |
| $et$ | Light-extinction coefficient | O’Flaherty 2019a | m | 0.26–0.31 |
| $H$ | Mean water-column depth | O’Flaherty 2019a | m | 0.5–6 |
| $V$ | Effective pool volume at water site | O’Flaherty 2019a | L | $6.75 \times 10^7$–<br>$8.25 \times 10^7$ |

### *E. coli* decay in soil

Empirical data on *E. coli* decay in litter-amended soil were obtained from Sharma Manan (Sharma Manan et al. 2019). Data consisted of log-transformed *E. coli* concentrations in the soil measured at several time points (days post-inoculation, *dpi*).

To characterize *E. coli* decline, an exponential decay function was assumed to follow:

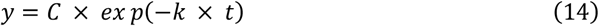

where y is the log-transformed *E. coli* concentration at time t (day), C is the initial concentration parameter, and k is the decay rate constant. The parameters C and k were estimated via nonlinear least squares:

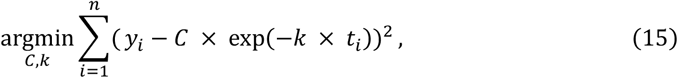

### Litter Application

The temporal evolution of the ESBL *E. coli* load on the soil was modeled for each Monte Carlo simulation i by a first-order decay of the initial litter-derived load. In our model, we therefore describe:

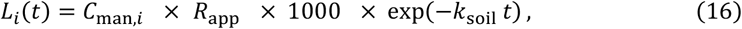

where *L*_*i*_(*t*) (CFU/m^2^) is the ESBL *E. coli* load at time *t* (days) after application, *C*_man,*i*_ (CFU. g−^1^) is the Monte Carlo draw of the litter concentration from the farm module output *C*_final_, *R*_app_ (kg/m^2^) is the application rate, the factor 1000 converts kilograms to grams, and *k*_soil_ is the decay constant.

### River Transport and Decay Model

We route the soil-derived load *L*_*i*_(*t*) into a simplified runoff and in-stream decay scheme, for each Monte Carlo iteration *i* and day *t*. Surface wash-off is computed once per day based on a constant fraction of freely mobile bacteria, then that wash-off load is instantly delivered to the receiving water body (i.e. travel time is assumed negligible on the daily time-step).

### Wash-off load

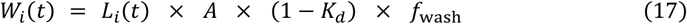

Here, *W*_*i*_(*t*) (CFU/day) is the total bacterial load washed off from the field on day *t* in simulation *i. L*_*i*_(*t*) (CFUm^2^) is the soil-module output, *A* (m^2^) is the field area, *K*_*d*_is the partition coefficient (fraction of bacteria sorbed to soil), and *f*_wash_ is the wash-off fraction of freely mobile bacteria. We compute wash-off once per day as a fixed fraction of the field load that is free for transport, representing the expected (event-averaged) daily export due to rainfall/runoff. By routing *W*_*i*_(*t*) directly into the river on the same day, we are effectively assuming that the farm-to-stream distance is short enough that travel time can be ignored at daily resolution.

### In-stream decay rate

Mancini’s Equation *(Mancini 1978)* was applied to capture in-river *E. coli* decay as a function of water temperature (T), salinity, and solar radiation. The daily first-order decay rate k was described as:

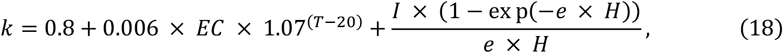

where *EC* is salinity (%), I is the solar radiation (ly/hr), e is the light extinction coefficient, and H is water column depth (m).

### Accumulated river load

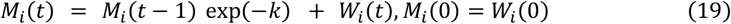

Where *M*_*i*_(*t*) (CFU) is the cumulative ESBL *E. coli* count in the water site on day *t*. Each day, the previous day’s count decays by exp (−*k*) and the new wash-off input *W*_*i*_(*t*)is added.

Dividing the accumulated *M*_*i*_(*t*) by the site volume *V* (L) yields *the* ESBL *E. coli* concentration *C*_*i*_(*t*)I n colony-forming units per liter:

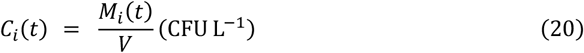

### Time notation (soil and river modules)

We define continuous time *t*as days since litter application, with *t* = 0 at application. Simulation outputs are stored and reported on discrete post-application days d *d* = 1,2, …, where day *d* represents the interval *t* ∈ (*d* − 1, *d*]. The soil module outputs loads *d* ≥ 1, the rive**r** module uses the same day index with zero initial load at *t* = 0 and updates concentrations from the day *d* export and in-stream decay.

### Lettuce module

To estimate *E. coli* contamination on lettuce, we linked daily irrigation-water concentrations *C*_*i*_(*t*) (CFU/mL) from the river model to leaf-surface adhesion and biphasic die-off on the crop (Table 3), following a previously proposed framework by O’Flaherty (O’Flaherty et al. 2019b). We evaluated scenarios with different intervals between land application of litter and lettuce planting. We define Δ*AP* as the days between application of litter and planting, with the environmental time origin at *t* = 0 on the day of application, thus planting occurs at *t* = Δ*AP*. In the lettuce module, irrigation begins on the planting day and proceeds daily through the growth period (35 days), so Δ*AP*is operationally equivalent to the interval from application to the first irrigation event. Leaf die-off is indexed on a leaf clock *τ* = 1 … 35.

**Table 3.**
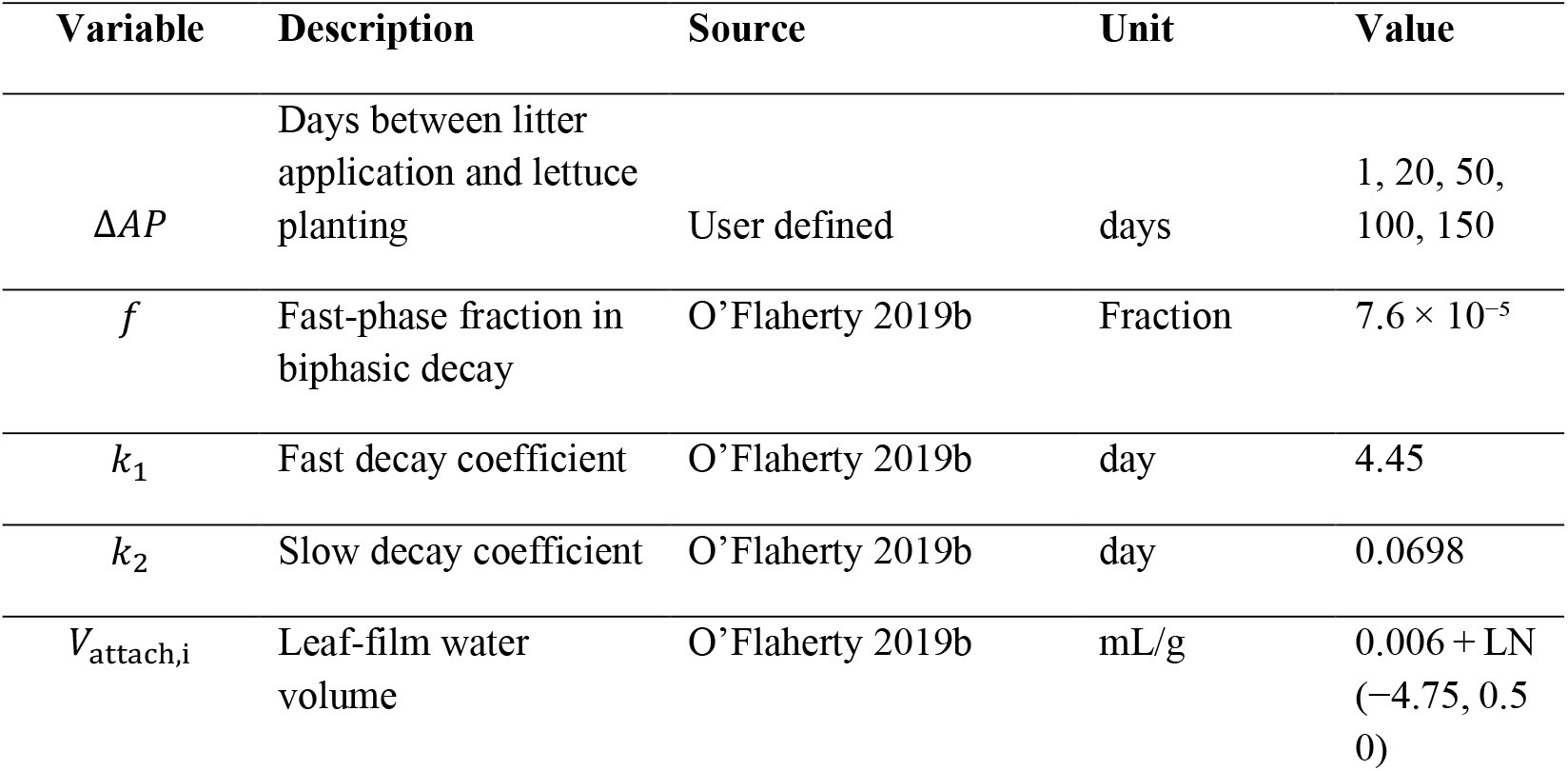
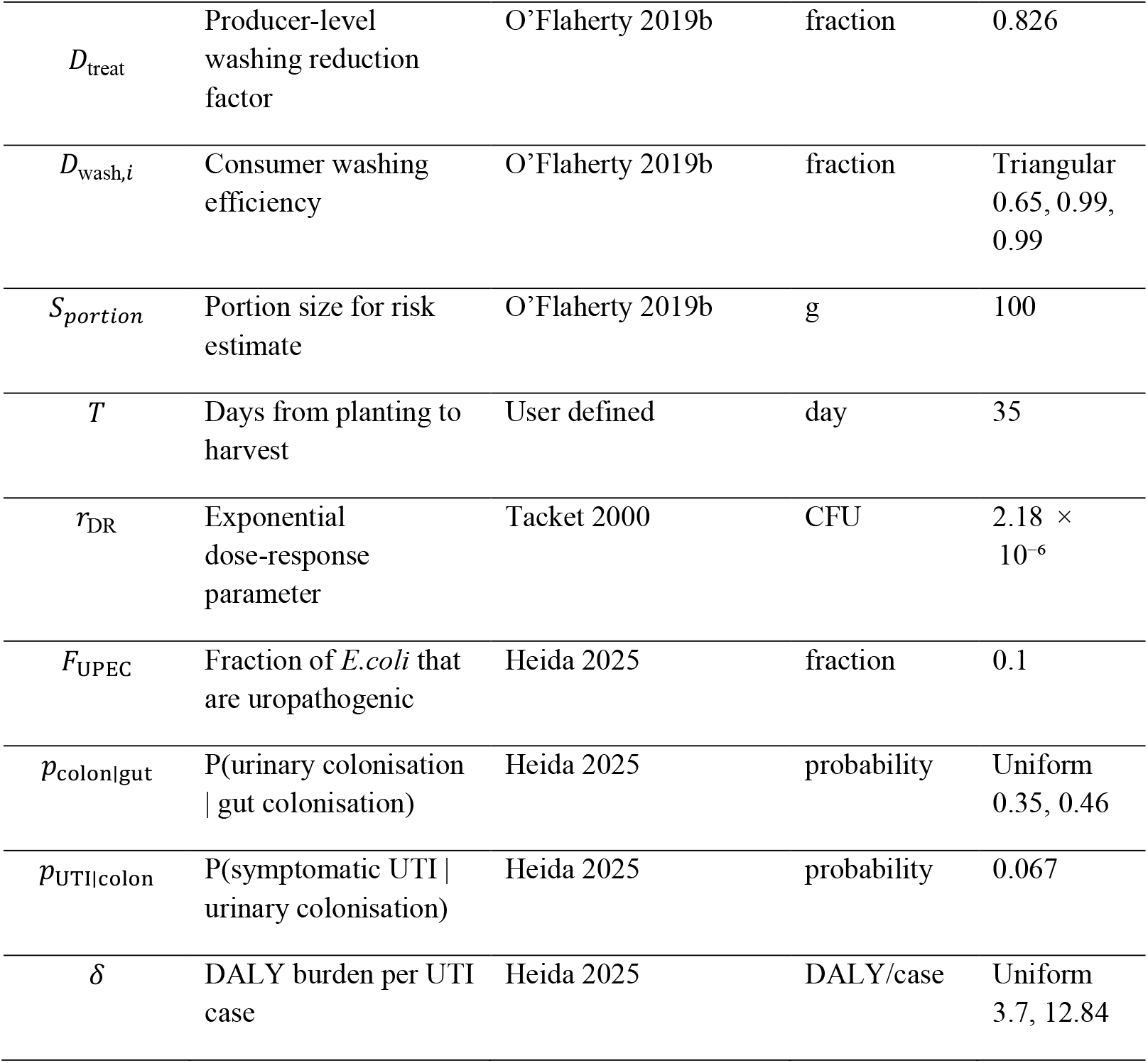
Key input variables for the lettuce module.

### Leaf-surface adhesion

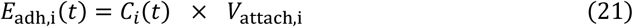

where *E*_adh,i_(*t*) (CFU/g) is the bacteria adhering to leaves, and *V*_attach,i_ (mL/g) is the water attachment volume.

### Biphasic decay on leaves

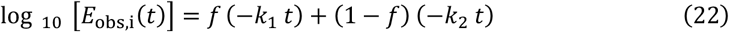

and

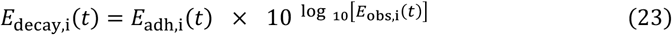

where a fraction *f* of cells decays at the fast rate *k*_1_ (/day) and the remainder (1 − *f*) at the slower rate *k*_2_ (/day). Here, *t* is days since first irrigation after planting.

### Accumulated contamination at harvest

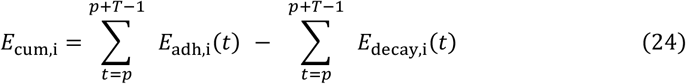

where *p* is the planting day, *T* is the growth duration (35 days), and the sums collect adhesion and decay over the growth period to compile the accumulated contamination at harvest *E*_cum,i_**Post-harvest washing**

A washing reduction factor *D*_treat_ represents the fraction of bacteria removed by producer washing with water. The concentration at harvest is calculated as:

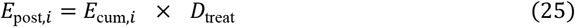

Furthermore, we modelled two scenarios: the consumption of 100g of lettuce *S*_*portion*_ by consumer of not further washed lettuce, as:

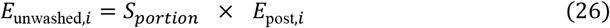

and the consumption of *S*_*portion*_ with an additional home washing (*D*_wash,*i*_), as:

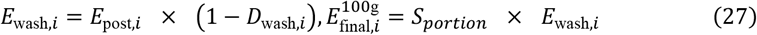

### Probability of infection and DALY estimation

We mapped lettuce dose (CFU per 100 g serving) to health outcomes using a pooled exponential dose-response for enteropathogenic *E. coli* (EPEC) with a fecal-shedding endpoint as a surrogate for gastrointestinal (GI) colonization. The parameterization ultimately traces to human volunteer challenge with EPEC O127:H6 (strain E2348/69); colonization in those studies was determined by recovery/shedding of the challenge strain from stools (primary source and trial details in Tacket et al., 2000). We adopted this surrogate because ESBL- or UPEC-specific human DR data are not available, using the approach taken by Heida et al., 2025 for ESBL *E. coli* in recreational water.

To translate GI colonization to UTI burden, we followed Heida’s framework: (i) apply a UPEC-like fraction to the ingested ESBL population; (ii) use conditional probabilities for urinary tract colonization given gut colonization and for symptomatic UTI given urinary colonization; and (iii) compute DALY per serving from outcome probabilities and published disability weights/durations.

The probability of GI is calculated as:

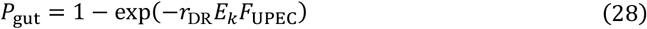

where *r*_DR_ is the exponential dose-response parameter,*E*_*k*_is the ingested dose (CFU) in a single event (lettuce consumption) and *F*_UPEC_ is the fraction of ESBL *E*.*coli* that are uropathogenic. Chicken feces (and thus litter) commonly contain *E. coli* populations in which a minority meet ExPEC criteria, and some of these are UPEC-like; published prevalences range from ∼13% ExPEC in broiler feces (with many categorized as UPEC) to ∼23% ExPEC in retail poultry meat. We therefore set *F*_UPEC_ = 0.10 as a realistic, conservative central, in accordance with Heida et al., 2025 (Stromberg et al. 2017; Mellata 2013; Lyhs et al. 2012; Literák et al. 2012).

The probability of UTI is estimated as:

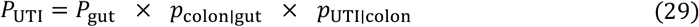

where *p*_colon|gut_ is the conditional probability of urinary tract colonization given gut colonization and *p*_UTI|colon_ is the conditional probability of symptomatic UTI given urinary tract colonization.

Finally, we estimated the DALY as:

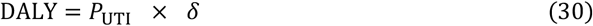

where *δ* is the DALY burden per UTI case (DALY/case).

### Sensitivity Analysis

We performed a variance-based sensitivity analysis using Partial Rank Correlation Coefficients (PRCC) to quantify how variability in the model inputs propagates to uncertainty in the litter load and in the final exposure outputs.

First, we identified key parameters spanning from each module, each varied uniformly over ±50 % of its baseline value. We generated 1000 stratified Latin Hypercube Sampling (LHS) draws across the n-dimensional input space, ensuring full coverage of each parameter’s marginal distribution without clustering.

For each of the 1000 parameter vectors, we executed the complete simulation chain, extracting the mean human exposure. Runs were parallelized with independent L’Ecuyer-Combined Multiple Recursive Generator substreams (L’Ecuyer, 1999).

Once the vector of output metrics was assembled, we computed PRCCs and derived 95 % confidence intervals through 100 bootstrap replicates. PRCC measures the monotonic relationship between each input and the outcome, controlling for all other parameters; those whose intervals excluded zero were deemed statistically significant.

### Software

All analyses and simulations were implemented in R (version 4.3.1) and RStudio (version 2024.09.1 build 394). Key packages including tidyverse (Wickham et al. 2019) for data manipulation and visualization, mc2d (Pouillot et al. 2016) and triangle (Carnell 2022) for distribution sampling, nls2 (Grothendieck 2013) for nonlinear regression, furrr (Bengtsson-Palme et al. 2023) for parallel computation, sensitivity (Iooss et al. 2024), and lhs (Carnell 2016) for sensitivity analysis. The full QMRA model is openly available on ENVIRE GitHub, https://github.com/ENVIRE-JPIAMR.

## Results and discussion

### Farm module

ESBL *E. coli* concentration in litter rose sharply after day 2 (16 × 10^-3^ CFU/g, SD 5 × 10^-4^, UI 7.6 × 10^-4^–2.7 × 10^-4^), reaching a maximum at day 8 (3.7 × 10^4^ CFU/g, SD 1.7 × 10^3^, UI 3.33–3.90 × 10^4^), before declining to 1.6 × 10^4^ CFU/g (SD 16.1, UI 1.6–1.60 x10^4^) at day 36 (Figure 2).

**Figure 2.**
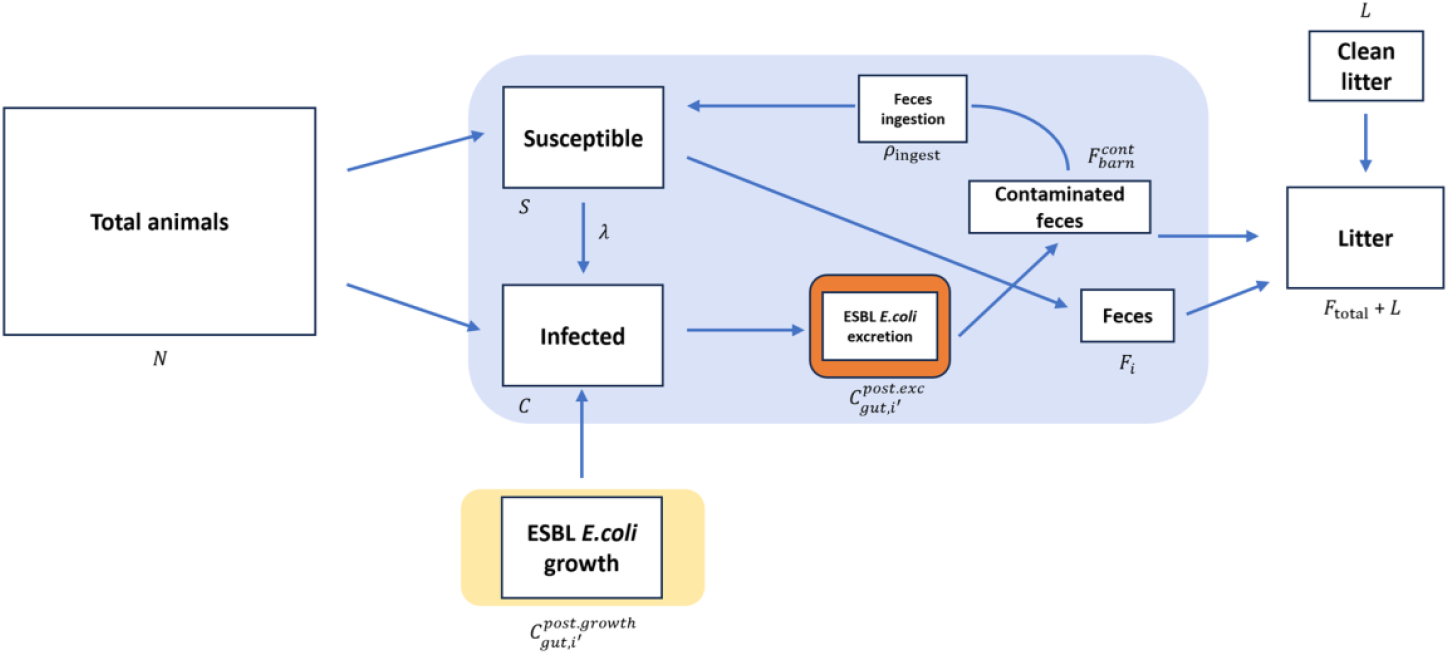
Diagram of the farm module simulating the transmission dynamics of ESBL-producing *E. coli* within a broiler flock.

**Figure 2.**
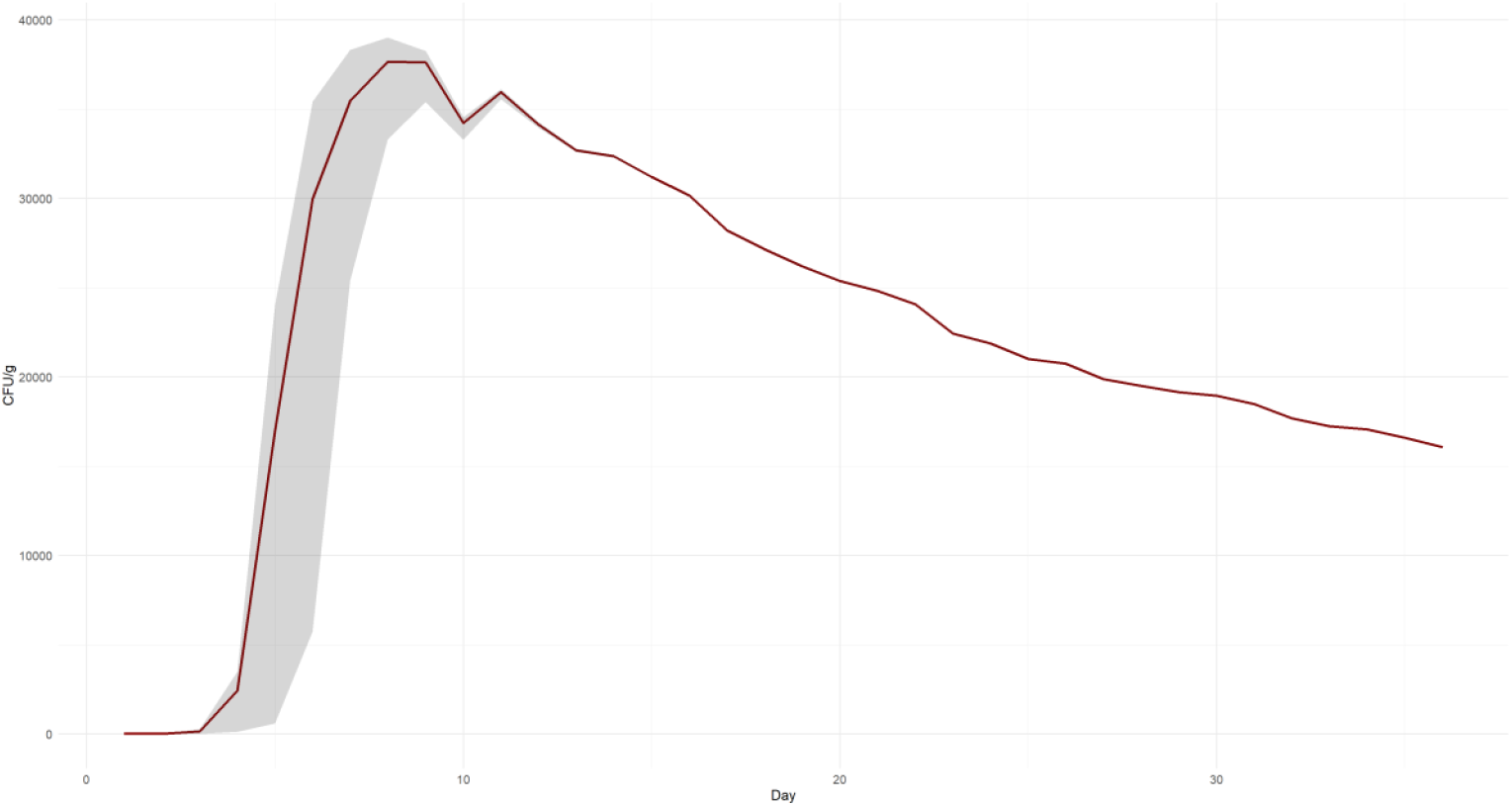
ESBL *E. coli* concentration (CFU/g) in litter over the broiler production period. Mean (dark red line) with 95 % uncertainty interval (shaded grey; 2.5th–97.5th percentiles) of 1000 iterations.

The proportion of colonized broilers climbed from 65% (UI 11–76%) on day 3 to more than 99.9 % by day 7 (UI 99–99%), remaining essentially constant thereafter (Figure 3).

**Figure 3.**
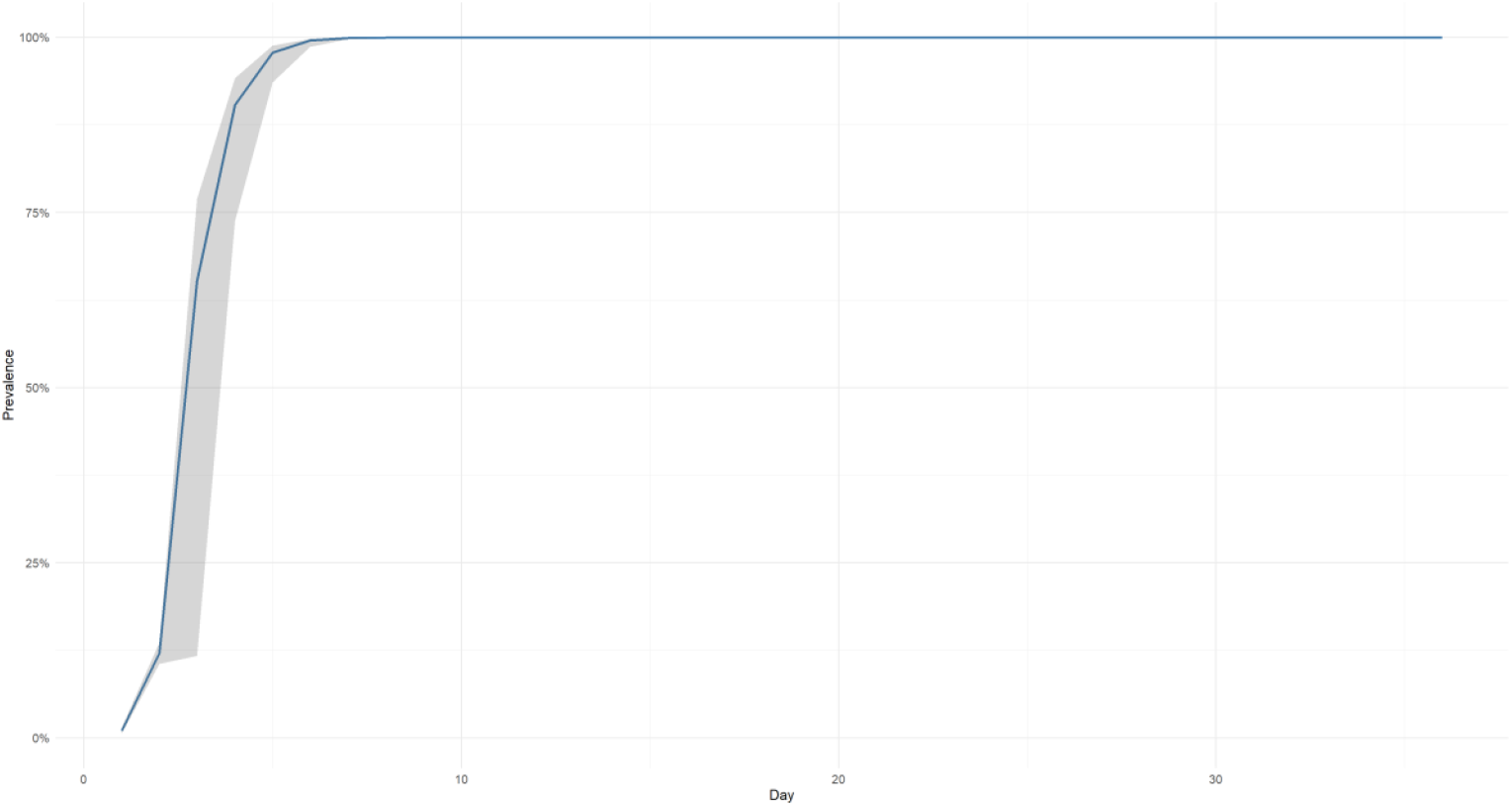
Prevalence of colonized broilers over time. Mean (blue line) with 95 % uncertainty interval (shaded grey; 2.5th–97.5th percentiles) of 1000 iterations.

The farm module simulates a rapid and efficient spread of ESBL *E. coli* within broiler flocks, consistent with field data and experimental studies (Ceccarelli Daniela et al. 2017; Kasparaviciene et al. 2025; Huijbers et al. 2016; Robé et al. 2021). In our simulation, flock-level colonization approached 100% within the first week. This pattern is similar to longitudinal studies where prevalence in day-old chicks often rises from negligible levels to near saturation within 5 to 7 days (Huijbers et al., 2018; Kasparaviciene et al., 2024; Dierikx et al., 2013). For example, Kasparaviciene et al. (2024) detected no ESBL *E. coli* on day 0, but found a 57.5% prevalence by day 5, while Huijbers et al. (2018) reported increases from 0-24% at day 1 to over 96% by day 7. These dynamics are biologically plausible given the extremely low infectious dose required for colonization, as few as 10 to 100 CFU can establish persistent colonization in broilers (Robé et al. 2019; Robé et al. 2021).

The concentration of ESBL *E. coli* in the litter estimated by this module aligns with the lower to mid-range of reported concentrations, though literature values vary widely. Tofani et al. (2022) measured ESBL/AmpC-*E. coli* loads in broiler cecal contents between 2.85 and 4.17 log10 CFU/g, while Atanasova et al. (2025) reported up to 5.48 log10 CFU/g in fresh broiler litter. At the lower end, Blaak et al. (2015) found a mean concentration of 2.4 × 104 CFU/kg in feces from Dutch farms. Also Becker et al. (2022) quantified a lower number of CFU/g in broiler litter. Such discrepancies are likely influenced by multiple factors including broiler age, diet, specific bacterial strains, litter conditions, sampling methods, and farm management. The model’s simplified assumption of logistic growth and a constant excretion rate does not fully capture transient increases in shedding that may occur due to stressors such as thinning, transport, or diet. As a result, the model may underestimate peak shedding under certain real-world scenarios.

Finally, our model underlines a critical message: once ESBL-producing *E. coli* is introduced in the barn, it spreads rapidly and is difficult to suppress (Robé et al. 2024; Luyckx et al. 2015). Because broiler colonization requires only a minimal infectious dose, even small biosecurity lapses at chick placement can result in widespread contamination.

In our simulation, we applied a Susceptible-Infected (colonized) (SI) transmission model, assuming that once a broiler becomes colonized it remains so until the end of the production cycle. Persistent colonization of gut bacteria like *E. coli* in broilers is well documented (Robé et al. 2019), yet this simplification may overlook more complex within-host dynamics (Galarneau et al. 2020). Dankittipong et al. (2023) compared both SI and Susceptible-Infectious-Susceptible (SIS) frameworks to study carbapenemase- and ESBL-producing *E. coli*, highlighting different transmission rates among resistant strains. Likewise, Becker et al. (2022) and Dame-Korevaar et al. (2020b) found the SI model adequate for capturing within-flock spread in broilers. By contrast, Furusawa et al. (2024) developed two compartmental models - one with a time-dependent decline in susceptibility and one with partial immunity to phylogenetic groups - to describe ESBL-producing *E. coli* transmission in Dutch broiler chains. Those models also incorporated environmental contamination between production cycles and within flocks, offering a more detailed framework for testing interventions and estimating public health risk.

Nevertheless, given the evidence for long-term carriage in broiler chickens, we judged the SI model the most consistent with our “worst case scenario” approach, since it reflects a permanent infection state once colonization occurs.

The sensitivity analysis of the farm module highlights three overwhelmingly dominant drivers of within-flock bacterial loads (Figure 4). First, the on-farm decay rate *ϕ* shows a very strong negative correlation with mean litter *E. coli* concentration (PRCC ≈ -0.98), indicating that small increases in bacterial die-off sharply reduce end-cycle loads. Conversely, gut carrying capacity *K* (PRCC ≈ +0.93) and the shedding rate *ϵ* (PRCC ≈ +0.93) both exhibited very strong positive correlations, underscoring that higher maximal gut loads and greater per-bacterium excretion rates drive litter contamination. Secondary factors include water metabolic loss *α* (PRCC ≈ -0.16) and target broiler weight *w*_tar_ (PRCC ≈ -0.04), which have moderate negative effects. All other parameters have PRCCs near zero with confidence intervals spanning zero, indicating negligible impact under the studied ranges. These results point to the critical importance of both bacterial die-off dynamics and host-level factors (gut capacity and shedding) in shaping ESBL *E. coli* loads on broiler farms.

**Figure 4.**
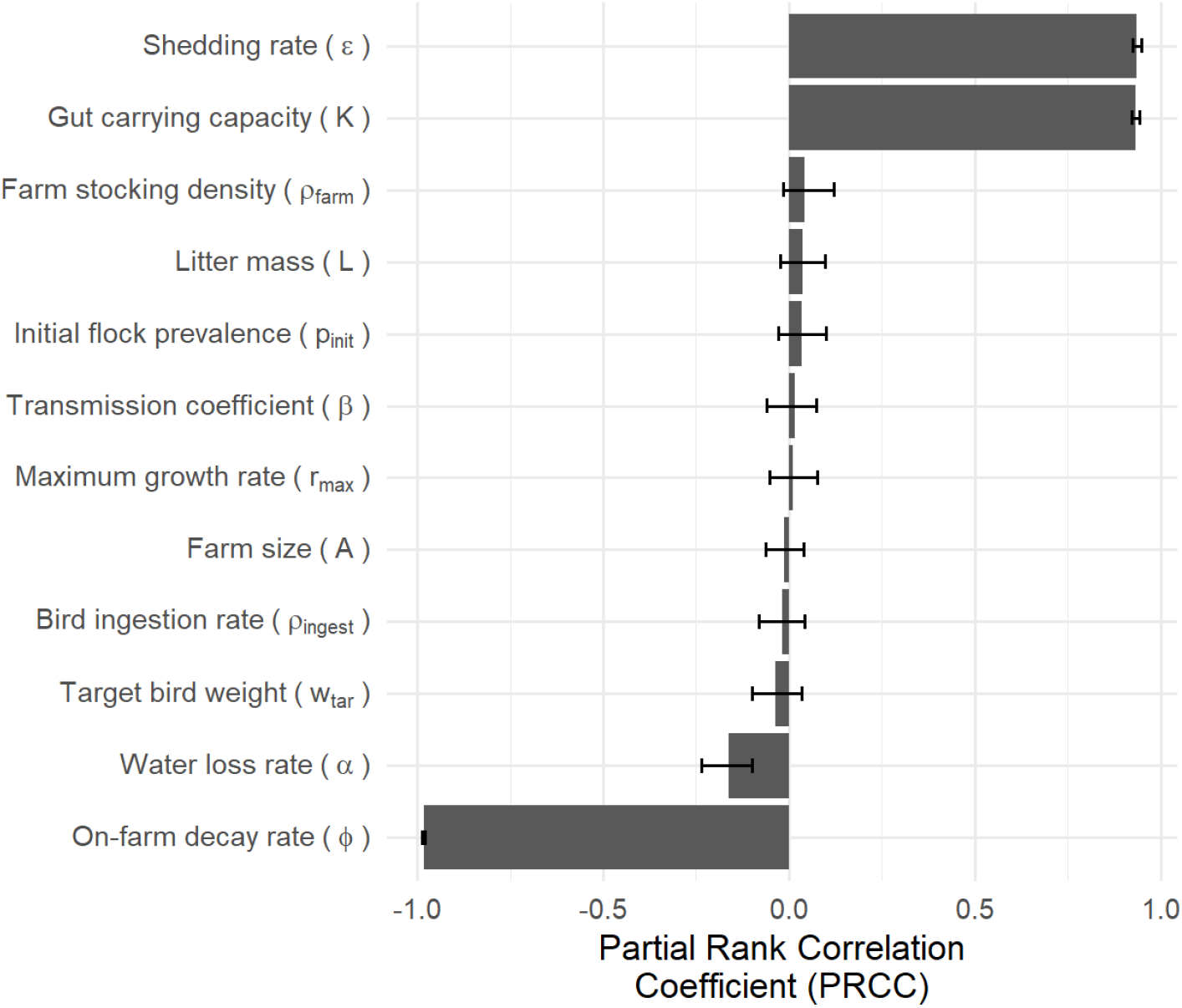
Sensitivity analysis litter ESBL *E. coli* load (CFU/g litter) at day 36; displayed is the mean Partial Rank Correlation Coefficient including 95% confidence intervals.

### Soil & River modules

When expressed per m^2^, the mean ESBL *E. coli* loads started at 3.2 × 10^7^ (SD 3.2 × 10^4^, UI 3.2– 3.2 × 10^7^) CFU/m^2^ on day 1 post litter application, fell to 2.2 × 10^7^ (SD 2.2 × 10^4^,UI 2.2–2.2 × 10^7^) CFU/m^2^ at day 10, and declined further to 8.6 × 10^5^ (SD 8.8 × 10^2^, UI 8.6–8.6 × 10^5^) CFU/m^2^ by day 100. This per-area view mirrors the per-mass decay and confirms that fields receive a substantial bacterial pulse immediately after litter application, but surface-area concentrations drop substantially within three months. Consequently, while broiler litter is a pronounced short-term source of soil contamination, its long-term contribution to human exposure via soil contact is minimal under baseline conditions. A limitation of our model is the lack of studies to use for validating the soil module. Most of the literature focuses on generic *E. coli* (Sarnino et al., 2025) or uses CFU/g of soil mixed with manure as quantitative output (Gekenidis et al. 2020). It is also important to specify that another big limitation of our model is the simplified litter application, as we assumed the ESBL *E. coli* to spread uniformly following a fixed application rate.

Simulated ESBL *E. coli* in river water decreased from 8.3 × 10^-2^ CFU/mL (SD 4.8 × 10^-3^, UI 7.6– 9.2 × 10^-2^) on day 1 to 6.0 × 10^-2^ CFU/mL (SD 3.5 × 10^-3^, UI 5.5–6.6 × 10^-2^) by day 10 and reached 6.2 × 10^-5^ CFU/mL (SD 3.6 × 10^-6^, UI 5.7–6.9 × 10^-5^) by day 200. Early declines were modest, reflecting dilution and decay; by day 200, concentrations were essentially zero. These magnitudes fall within reported bathing-water ranges for AR/ESBL *E. coli* (predicted 0.45–345.09 CFU/100 mL as summarized by O’Flaherty et al., 2019a) and are consistent with ESBL fractions (0.05–1%) applied to recreational criteria (≈0.06–4.1 CFU/100 mL) in Heida et al., 2025.

### Lettuce module

Our model simulated the contamination of lettuce via irrigation with water containing ESBL *E. coli*, predicting contamination levels at harvest influenced by river contamination, application practices, and post-irrigation environmental factors, with risk reduction achieved through standard household washing. In our simulation, we estimated the exposure via ingestion of both washed by final consumers and unwashed lettuce. Furthermore, we compared harvest-time exposure across various intervals between litter application and lettuce planting. With a one-day interval, mean exposure reached 1.7 CFU/100 g (SD 8.5 × 10^-2^, UI 1.5–1.8) for unwashed lettuce and 0.2 CFU/100g (SD 0.1, UI 0.03–0.49) after a consumer wash, indicating that even simple rinsing may reduce loads by nearly 90 % (Figure 5). As the interval lengthened, exposures fell markedly: at 20 days, 0.85 (SD 5.0 × 10^-2^, UI 0.76–0.95) (unwashed) and 0.1 (SD 7.4 × 10^-2^, UI 0.01 – 0.25) (washed); at 150 days, 7.6 × 10^-3^ (SD 4.5 × 10^-4^, 6.7–8.6 × 10^-3^) CFU and 8.6 × 10^-4^ (SD 5.5 × 10^-4^, 1.1–2.0 × 10^-4^) CFU, respectively.

**Figure 5.**
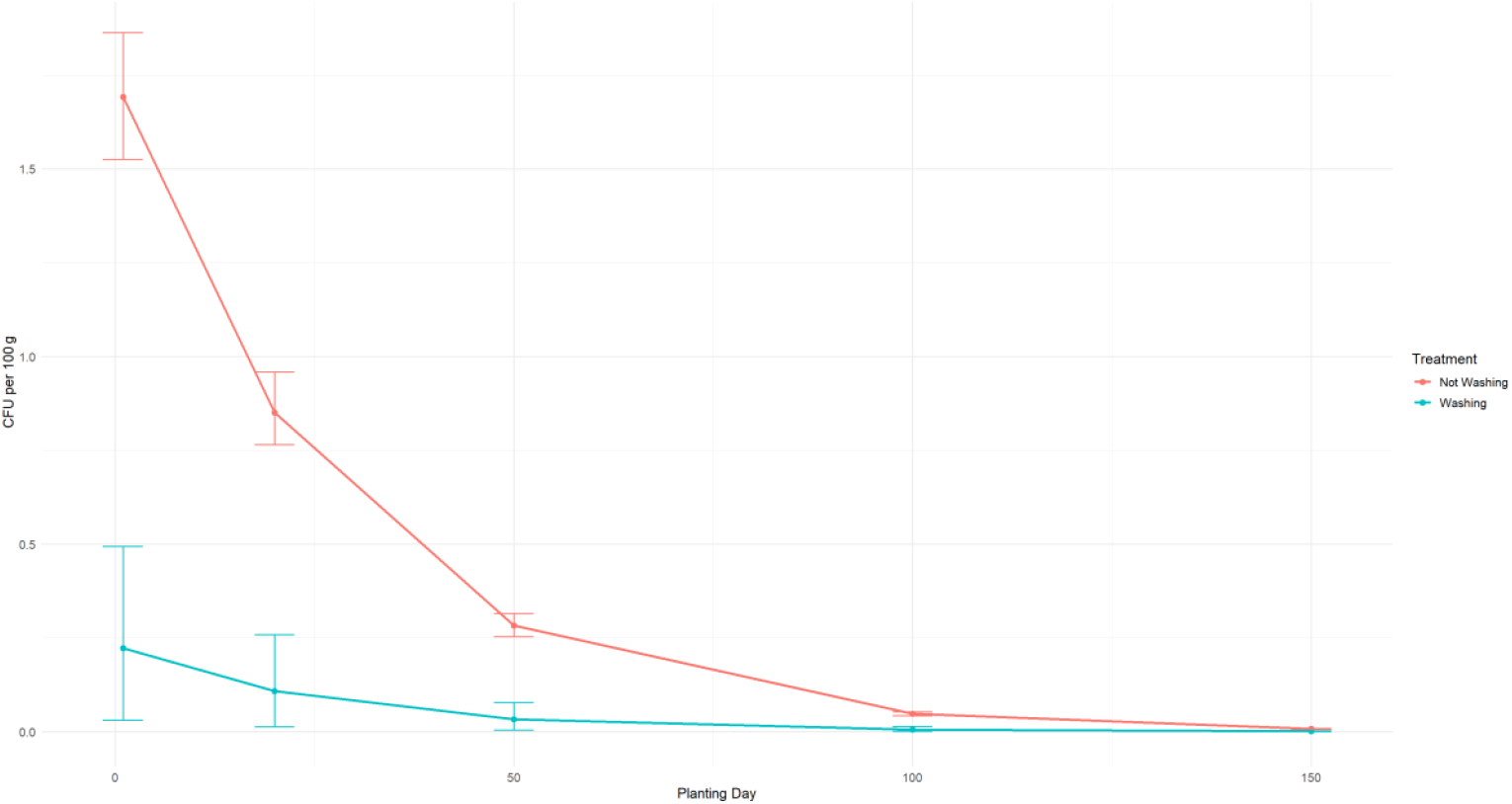
Predicted ESBL *E. coli* on lettuce at harvest (CFU/100 g) at different discrete intervals between broiler litter application and planting day. Lines & points show mean exposure; error bars show the 95 % uncertainty interval (2.5th–97.5th percentiles) across 1000 iterations.

Harvested lettuce doses were converted into infection probabilities and DALYs. Without washing, mean gut-colonization risks ranged from 3.30 × 10^-7^ on planting interval 1, corresponding to UTI risks of 9.02 × 10^-9^ and DALYs lost per serving of 7.48 × 10^-8^ (Table 4). Consumer washing and increasing the interval between litter application and lettuce planting reduced colonization risks, UTI risks, and DALYs. It appears that the rapid natural decay of *E. coli* on foliage and the incremental benefit of simple post-harvest washing reduce the risk of UTI and DALY burden from raw-leaf consumption. Our results are consistent with studies identifying irrigation water as a plausible route for pre-harvest contamination of fresh produce. However, we did not model or compare alternative routes and therefore do not rank their relative importance (Solomon et al.; Jensen et al. 2013; Holvoet et al. 2013; Iwu and Okoh 2019). Several studies have shown that *E. coli*, including pathogenic strains, can attach to and survive on lettuce surfaces for extended periods (Islam et al. 2004; Ingham et al. 2004). Environmental factors such as higher temperatures and light intensity tend to speed inactivation on plant surfaces (Weller et al. 2017; Ottoson et al. 2011). In our model, increasing the interval between litter application and planting reduces predicted contamination. This is consistent with studies showing time-dependent decline of *E. coli* in manure-amended soils and reduced detection on crops with longer intervals after application (Ingham et al. 2005; Black et al. 2021).

**Table 4.** Mean and UI (in brackets) probability of gastrointestinal colonization (GI) and urinary tract infection (UTI), and corresponding DALY per exposure event for each planting interval and exposure scenario (washed vs. unwashed lettuce).

| Planting Interval | Risk GI | Risk UTI | DALY | Exposure |
| --- | --- | --- | --- | --- |
| 1 | $3.3 \times 10^{-7}$ ($2.9 \times 10^{-7}$–$3.6 \times 10^{-7}$) | $9.0 \times 10^{-9}$ ($7.5 \times 10^{-9}$–$1.0 \times 10^{-8}$) | $7.5 \times 10^{-8}$ ($3.4 \times 10^{-8}$–$1.2 \times 10^{-7}$) | Unwashed |
| 1 | $4.3 \times 10^{-8}$ ($6.0 \times 10^{-9}$–$9.6 \times 10^{-8}$) | $1.1 \times 10^{-9}$ ($1.6 \times 10^{-10}$–$2.6 \times 10^{-9}$) | $1.0 \times 10^{-8}$ ($1.2 \times 10^{-9}$–$3.0 \times 10^{-8}$) | Washed |
| 20 | $1.6 \times 10^{-7}$ ($1.5 \times 10^{-7}$–$1.8 \times 10^{-7}$) | $4.5 \times 10^{-9}$ ($3.6 \times 10^{-9}$–$5.2 \times 10^{-9}$) | $3.6 \times 10^{-8}$ ($1.7 \times 10^{-8}$–$5.8 \times 10^{-8}$) | Unwashed |
| 20 | $2.1 \times 10^{-8}$ ($2.6 \times 10^{-9}$–$5.0 \times 10^{-8}$) | $5.8 \times 10^{-10}$ ($7.1 \times 10^{-11}$–$1.3 \times 10^{-9}$) | $4.7 \times 10^{-9}$ ($4.9 \times 10^{-10}$–$1.3 \times 10^{-8}$) | Washed |
| 50 | $5.5 \times 10^{-8}$ ($4.9 \times 10^{-8}$–$6.1 \times 10^{-8}$) | $1.5 \times 10^{-9}$ ($1.2 \times 10^{-9}$–$1.8 \times 10^{-9}$) | $1.1 \times 10^{-8}$ ($5.6 \times 10^{-9}$–$1.8 \times 10^{-8}$) | Unwashed |
| 50 | $6.5 \times 10^{-9}$ ($8.2 \times 10^{-10}$–$1.5 \times 10^{-8}$) | $1.7 \times 10^{-10}$ ($2.2 \times 10^{-11}$–$4.3 \times 10^{-10}$) | $1.5 \times 10^{-9}$ ($1.6 \times 10^{-10}$–$3.9 \times 10^{-9}$) | Washed |
| 100 | $9.3 \times 10^{-9}$ ($8.3 \times 10^{-9}$–$1.0 \times 10^{-8}$) | $2.5 \times 10^{-10}$ ($2.1 \times 10^{-10}$–$3.0 \times 10^{-10}$) | $2.2 \times 10^{-9}$ ($9.0 \times 10^{-10}$–$3.4 \times 10^{-9}$) | Unwashed |
| 100 | $1.1 \times 10^{-9}$ ($1.5 \times 10^{-10}$–$2.8 \times 10^{-9}$) | $3.1 \times 10^{-11}$ ($3.6 \times 10^{-12}$–$7.8 \times 10^{-11}$) | $2.3 \times 10^{-10}$ ($2.2 \times 10^{-11}$–$5.4 \times 10^{-10}$) | Washed |
| 150 | $1.5 \times 10^{-9}$ ($1.3 \times 10^{-9}$–$1.6 \times 10^{-9}$) | $4.0 \times 10^{-11}$ ($3.4 \times 10^{-11}$–$4.9 \times 10^{-11}$) | $3.2 \times 10^{-10}$ ($1.5 \times 10^{-10}$–$5.1 \times 10^{-10}$) | Unwashed |
| 150 | $1.7 \times 10^{-10}$ ($2.2 \times 10^{-11}$–$3.9 \times 10^{-10}$) | $4.6 \times 10^{-12}$ ($5.6 \times 10^{-13}$–$1.0 \times 10^{-11}$) | $3.7 \times 10^{-11}$ ($4.2 \times 10^{-12}$–$1.0 \times 10^{-10}$) | Washed |

Regarding post-harvest mitigation at the consumer level, the model prediction of limited efficacy on *E. coli* complete reduction for household washing is strongly supported by experimental data (Uhlig et al. 2017; Jensen et al. 2015; Lopez et al. 2015). However, properly rinsing green leaves may reduce the consumer’s risk (Ottoson et al. 2011). The limitations of consumer-level washing practices firmly place the primary responsibility for ensuring the microbial safety of fresh produce, particularly regarding contaminants introduced via irrigation, on upstream controls within the agricultural production system.

The sensitivity analysis (Figure 6) showed that lettuce-borne ESBL *E. coli* exposure is overwhelmingly governed by how readily bacteria sorb to soil particles: the partition coefficient has a very strong negative association (PRCC ≈ -0.90), meaning that greater sorption reduces transfer to runoff and, ultimately, to foliage (Sowah et al., 2020). Soil decay kinetics also matter: the estimated decay rate in soil and the environmental decay rate on the farm both correlate negatively (PRCC ≈ -0.36 and -0.31, respectively), indicating that faster bacteria die-off sharply cuts lettuce contamination. On the flip side, parameters that increase bacterial availability such as wash-off fraction from soil to water (PRCC ≈ +0.17) and the shedding rate in the farm module (PRCC ≈ +0.17) modestly raise exposure. Litter mass (kg/m^2^) and gut carrying capacity also exert smaller but significant influences. Together, these results suggest that strategies aiming to enhance soil-borne decay and reduce bacterial release during runoff will be most effective at reducing ESBL *E. coli* levels on harvested lettuce.

**Figure 6.**
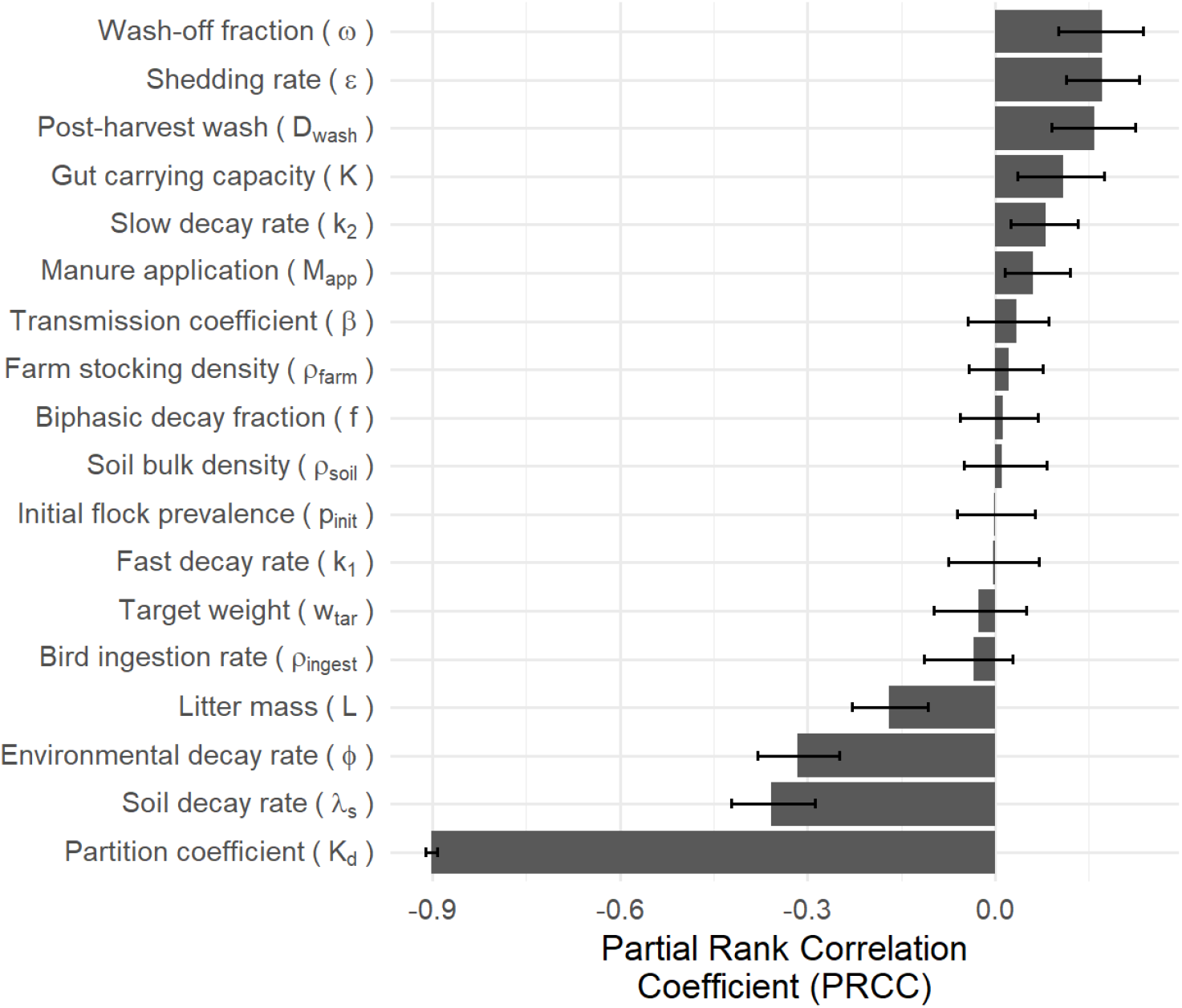
Sensitivity analysis for lettuce consumption (single event). Displayed is the mean Partial Rank Correlation Coefficient including 95% confidence intervals.

### Limitations

Our model incorporates several simplifying assumptions that merit further refinement. In general, our model represents a worst-case scenario, potentially overestimating human exposure. First, in the farm module, we treat *E. coli* as uniformly distributed across all fecal matter and assume each broiler remains colonized from the moment of infection until cycle end, with bacterial growth governed by a single logistic curve. In reality, fecal deposition is spatially clustered, broiler immune responses and stress levels vary over time, and diversity among ESBL *E. coli* strains can differ, factors that can drive transient “hot spots” of shedding and alter transmission dynamics.

Second, the soil module idealizes litter spreading as perfectly homogeneous and represents die-off with one constant exponential rate. This ignores the influence of temperature fluctuations, soil moisture, pH gradients, bioturbation by earthworms and other fauna, and incorporation by tillage all of which can create microsites where bacteria persist or die off more rapidly. Furthermore, litter is assumed to be spread on fields immediately after flock removal (0 days storage) and without any treatment (such as composting), ensuring maximal viable ESBL *E. coli* loads at application. Nevertheless, although manure treatment and storage are common practices (Sarnino et al., 2025), the Nitrates Directive (91/676/EEC), the main EU legislation regulating manure use to protect water quality, does not mandate treatments or set a waiting period.

Third, our river transport component applies fixed wash-off fractions and partition coefficients, yielding a smooth decline in contaminant load. Yet real watersheds undergo storm-driven pulses, variable flow regimes, sediment resuspension, channel morphology effects, and spatial heterogeneity in land cover, which together can produce episodic peaks in bacterial concentrations that our model may miss. As a potential improvement, incorporation of a full SWAT model could better capture these complex hydrological and land-use dynamics (Douglas-Mankin et al, 2010).

Fourth, the lettuce module relies on a biphasic decay curve with uniform attachment efficiency to leaf surfaces and a single removal efficiency for post-harvest washing. It does not account for differences in leaf microstructure, irrigation method (e.g., overhead versus drip), spray droplet size, canopy microclimate, or commercial-scale wash protocols.

Furthermore, our framework omits key microbial ecology processes, most notably horizontal transfer of resistance genes (e.g. via plasmids) among environmental bacteria and interactions with other microbial populations that may inhibit or promote ESBL *E. coli* survival. We also do not capture potential regrowth phases under favorable conditions.

Another dominant uncertainty is the soil-water attachment/partitioning of ESBL *E. coli* under local conditions-i.e., the proportion that is particle-associated versus freely suspended. Ideal data would quantify, after rainfall or irrigation in litter-amended plots, bacteria in the water phase and in the sediment phase, or use simple laboratory tests to estimate the particle-association fraction for local soils.

Our dose-response model maps from an EPEC human challenge model (fecal shedding as a proxy for GI colonization) to UTI via conditional transitions and a UPEC-like fraction. This surrogate is necessary because no ESBL- or UPEC-specific human DR exists; consequently, risk outputs should be interpreted as scenario-based indicators with uncertainty concentrated in the UPEC fraction and conditional transitions rather than as clinical predictions. Heida et al. 2025 emphasized the same caveats for recreational waters; we adopt that caution here. Evidence to date documents contamination of raw vegetables with ESBL/ExPEC *E. coli* but direct epidemiologic links between raw-vegetable consumption and UTI are limited (Balbuena-Alonso et al. 2023; Song et al. 2020).The biological plausibility rests on GI colonization serving as a reservoir for UPEC and periurethral transfer, with sex differences in UTI incidence not resolved in our per-serving DALY (Josephs-Spaulding et al. 2021).

Finally, to our knowledge, this is the first ESBL *E. coli* focused QMRA connecting broiler flock, litter-amended soil, receiving water and lettuce consumption in a single framework. For this reason, a head-to-head validation against a prior end-to-end model is not yet possible. We therefore adopted a modular validation strategy and, where feasible, compared predicted magnitudes and expected signatures with published models and experimental studies.

Despite these simplifications, the model captures the dominant processes governing ESBL *E. coli* dissemination to produce and, by intentionally adopting conservative assumptions, provides exposure estimates that likely bound real-world outcomes. Consequently, we believe the results remain sufficiently robust to inform risk management and guide intervention strategies.

Future model iterations should introduce spatially explicit litter application, temperature and moisture driven decay rates, event-based hydrologic inputs, mechanistic leaf-surface colonization dynamics, and microbial community interactions, including gene transfer dynamics. Incorporating these elements will enhance realism, reduce uncertainty, and yield more robust estimates of human exposure risk.

## Conclusions

In this study, we developed and applied a four-module QMRA model to trace the journey of ESBL *E. coli* from broiler flocks through soil, surface water, and fresh produce. The farm module confirmed rapid amplification in litter, with near-complete flock colonization within one week and end-cycle litter loads in the low 10^4^ CFU/g range-values. Soil levels of ESBL *E. coli* peaked at about 3.2 × 10^7^ CFU/m^2^ right after spreading, then fell steadily and were substantially lower three months later. River concentrations showed a similar drop, starting near 8.3 × 10^-2^ CFU/mL on day 1, falling to 6.0 × 10^-2^ CFU/mL by day 10, and reaching under 10^-4^ CFU/mL by day 200. Lettuce simulations showed harvest loads of 1.7 CFU/100 g at one day interval between application of litter and planting, dropping to 0.85 CFU/100 g at 20 days, with simple household washing cutting exposure by ∼90 percent. Overall, in the lettuce module, the mean gut-colonization probabilities per 100 g serving did not exceed 10^-7^, UTI risks 10^-9^, and associated DALYs per serving 10^-8^, with household washing reducing risk levels. Sensitivity analyses identified the soil-water partition coefficient and decay rates as the most influential parameters, pointing to targeted data collection needs. Although natural decay, dilution, and interventions such as composting can reduce human exposure by orders of magnitude, persistent contamination calls for robust manure management and strict irrigation-water quality controls. Future work should introduce spatial heterogeneity in litter spreading, dynamic weather drivers, host immunity and horizontal gene transfer to refine risk estimates and guide effective on-farm and pre-harvest interventions.

## Data Availability

All data produced in the present work are contained in the manuscript

## Acknowledgments

The authors would like to thank all the members of the ENVIRE project consortium for their support.

## Author Contributions

NS: Writing - original draft, Formal analysis, Conceptualization, Visualization, Writing – review & editing. SB: Formal analysis, Conceptualization, Writing - review & editing. LC: Conceptualization, Writing - review & editing, Supervision. RM: Conceptualization, Writing - review & editing, Supervision, Funding acquisition.

## Supplementary data

The spreadsheet “Sarnino-et-al-2025b-results.xlsx” containing detailed simulation outputs is available online alongside this article.

## Publication bibliography

Agersø, Yvonne; Jensen Jacob Dyring; Hasman, Henrik; Pedersen, Karl (2014): Spread of Extended Spectrum Cephalosporinase-Producing Escherichia coli Clones and Plasmids from Parent Animals to Broilers and to Broiler Meat in a Production Without Use of Cephalosporins. In Foodborne Pathogens and Disease 11 (9), pp. 740–746. doi: 10.1089/fpd.2014.1742.

Alegbeleye, Oluwadara Oluwaseun; Sant’Ana Anderson S. (2020): Manure-borne pathogens as an important source of water contamination: An update on the dynamics of pathogen survival/transport as well as practical risk mitigation strategies. In International journal of hygiene and environmental health 227, p. 113524. doi: 10.1016/j.ijheh.2020.113524.

Apostolakos, Ilias; Mughini-Gras, Lapo; Fasolato, Luca; Piccirillo, Alessandra (2019): Assessing the occurrence and transfer dynamics of ESBL/pAmpC-producing Escherichia coli across the broiler production pyramid. In PLOS ONE 14 (5), e0217174. doi: 10.1371/journal.pone.0217174.

Atanasova, Aleksandra; Amon, Thomas; Friese, Anika; Rösler, Uwe; Merle, Roswitha; Herrmann, Christiane et al. (2025): Effects of Carbon–to–Nitrogen Ratio and Temperature on the Survival of Antibiotic-Resistant and Non-Resistant Escherichia coli During Chicken Manure Anaerobic Digestion. In Poultry 4, p. 9. doi: 10.3390/poultry4010009.

Balbuena-Alonso, Maria G.; Camps, Manel; Cortés-Cortés, Gerardo; Carreón-León, Eder A.; Lozano-Zarain, Patricia; Del Rocha-Gracia, Rosa Carmen (2023): Strain belonging to an emerging, virulent sublineage of ST131 Escherichia coli isolated in fresh spinach, suggesting that ST131 may be transmissible through agricultural products. In Frontiers in Cellular and Infection Microbiology Volume 13 - 2023. Available online at https://www.frontiersin.org/journals/cellular-and-infection-microbiology/articles/10.3389/fcimb.2023.1237725.

Becker, Evelyne; Correia-Carreira, Guido; Projahn, Michaela; Käsbohrer, Annemarie (2022): Modeling the Impact of Management Changes on the Infection Dynamics of Extended-Spectrum Beta-Lactamase-Producing Escherichia coli in the Broiler Production. In 2076-2607 10 (5). doi: 10.3390/microorganisms10050981.

Bengtsson-Palme, Johan; Abramova, Anna; Berendonk Thomas U.; Coelho Luis Pedro; Forslund Sofia K.; Gschwind Rémi et al. (2023): Towards monitoring of antimicrobial resistance in the environment: For what reasons, how to implement it, and what are the data needs? In 0160-4120 178, p. 108089. doi: 10.1016/j.envint.2023.108089.

Blaak, Hetty; van Hoek Angela H A M; Hamidjaja Raditijo A.; van der Plaats Rozemarijn Q J; Kerkhof-de Heer, Lianne; de Roda Husman, Ana Maria; Schets Franciska M. (2015): Distribution, Numbers, and Diversity of ESBL-Producing E. coli in the Poultry Farm Environment. In PLOS ONE 10 (8), e0135402. doi: 10.1371/journal.pone.0135402.

Black, Zoe; Balta, Igori; Black, Lisa; Naughton Patrick J.; Dooley James S. G.; Corcionivoschi, Nicolae (2021): The Fate of Foodborne Pathogens in Manure Treated Soil. In 1664-302X 12. Available online at https://www.frontiersin.org/journals/microbiology/articles/10.3389/fmicb.2021.781357.

Carnell, R. (2022): triangle: Distribution Functions and Parameter Estimates for the Triangle Distribution. In R package version 1.

Carnell, Rob (2016): Package ‘lhs’. In CRAN. https://cran.rproject.org/web/packages/lhs/lhs.pdf.

Ceccarelli Daniela; van Essen-Zandbergen Alieda; Smid Bregtje; Veldman Kees T.; Boender Gert Jan; Fischer Egil A.J. et al. (2017): Competitive Exclusion Reduces Transmission and Excretion of Extended-Spectrum-β-Lactamase-Producing Escherichia coli in Broilers. In Applied and Environmental Microbiology 83 (11), e03439–16. doi: 10.1128/AEM.03439-16.

Çekiç Samantha K.; De, Jaysankar; Jubair, Mohammad; Schneider Keith R. (2017): Persistence of Indigenous Escherichia coli in Raw Bovine Manure-Amended Soil. In 0362-028X 80 (9), pp. 1562–1573. doi: 10.4315/0362-028X.JFP-17-033.

Chantziaras, Ilias; Boyen, Filip; Callens, Bénédicte; Dewulf, Jeroen (2014): Correlation between veterinary antimicrobial use and antimicrobial resistance in food-producing animals: a report on seven countries. In J Antimicrob Chemother 69 (3), pp. 827–834. doi: 10.1093/jac/dkt443.

Chapman, B.; Pintar, K.; Smith, B. A. (2018): Multi-Exposure Pathway Model to Compare Escherichia coli O157 Risks and Interventions. In 0272-4332 38 (2), pp. 392–409. doi: 10.1111/risa.12826.

Collineau, Lucie; Chapman, Brennan; Bao, Xu; Sivapathasundaram, Branavan; Carson Carolee A.; Fazil, Aamir et al. (2020): A farm-to-fork quantitative risk assessment model for Salmonella Heidelberg resistant to third-generation cephalosporins in broiler chickens in Canada. In International Journal of Food Microbiology 330, p. 108559. doi: 10.1016/j.ijfoodmicro.2020.108559.

Dame-Korevaar, Anita; Fischer Egil A.J.; van der Goot, Jeanet; Velkers, Francisca; Ceccarelli, Daniela; Mevius, Dik; Stegeman, Arjan (2020a): Early life supply of competitive exclusion products reduces colonization of extended spectrum beta-lactamase-producing Escherichia coli in broilers. In 0032-5791 99 (8), pp. 4052–4064. doi: 10.1016/j.psj.2020.04.025.

Dame-Korevaar, Anita; Kers Jannigje G.; Goot, Jeanet; Velkers, Francisca; Ceccarelli, Daniela; Mevius, Dik et al. (2020b): Competitive Exclusion Prevents Colonization and Compartmentalization Reduces Transmission of ESBL-Producing Escherichia coli in Broilers. In Frontiers in Microbiology 11. doi: 10.3389/fmicb.2020.566619.

Dankittipong, Natcha; Alderliesten Jesse B.; van den Broek, Jan; Dame-Korevaar, M. Anita; Brouwer Michael S.M.; Velkers Francisca C. et al.(2023): Comparing the transmission of carbapenemase-producing and extended-spectrum beta-lactamase-producing Escherichia coli between broiler chickens. In Preventive Veterinary Medicine 219, p. 105998. doi: 10.1016/j.prevetmed.2023.105998.

Dierikx, Cindy M.; van der Goot Jeanet A.; Smith Hilde E.; Kant, Arie; Mevius Dik J. (2013): Presence of ESBL/AmpC -Producing Escherichia coli in the Broiler Production Pyramid: A Descriptive Study. In PLOS ONE 8 (11), e79005. doi: 10.1371/journal.pone.0079005.

Douglas-Mankin, K. R.; Srinivasan, Raghavan; Arnold, J. G. (2010): Soil and Water Assessment Tool (SWAT) model: Current developments and applications. In Transactions of the ASABE 53 (5), pp. 1423–1431.

Fastl, Christina; Carvalho Ferreira, Helena C. de; Babo Martins, Sara; Sucena Afonso, João; Di Bari, Carlotta; Venkateswaran, Narmada et al. (2023): Animal sources of antimicrobial-resistant bacterial infections in humans: a systematic review. In 0950-2688 151, e143. doi: 10.1017/S0950268823001309.

Furusawa, Minori; Widgren, Stefan; Evers Eric G.; Fischer Egil A.J. (2024): Quantifying health risks from ESBL-producing Escherichia coli in Dutch broiler production chains and potential interventions using compartmental models. In Preventive Veterinary Medicine 224, p. 106121. doi: 10.1016/j.prevetmed.2024.106121.

Galarneau, Karen D.; Singer Randall S.; Wills Robert W. (2020): A system dynamics model for disease management in poultry production. In 0032-5791 99 (11), pp. 5547–5559. doi: 10.1016/j.psj.2020.08.011.

Gekenidis, Maria-Theresia; Rigotti, Serena; Hummerjohann, Jörg; Walsh, Fiona; Drissner, David (2020): Long-Term Persistence of blaCTX-M-15 in Soil and Lettuce after Introducing Extended-Spectrum β-Lactamase (ESBL)-Producing Escherichia coli via Manure or Water. In 2076-2607 8 (11). doi: 10.3390/microorganisms8111646.

Grothendieck, Maintainer G. (2013): Package ‘nls2’. In Non-linear regression with brute force.

Heida, Ashley; Hamilton Mark T.; Gambino, Julia; Sanderson, Kaylee; Schoen Mary E.; Jahne Michael A. et al.(2025): Population Ecology-Quantitative Microbial Risk Assessment (QMRA) Model for Antibiotic-Resistant and Susceptible E. coli in Recreational Water. In 0013-936X 59 (9), pp. 4266–4281. doi: 10.1021/acs.est.4c07248.

Holvoet, Kevin; Sampers, Imca; Callens, Bénédicte; Dewulf, Jeroen; Uyttendaele, Mieke (2013): Moderate Prevalence of Antimicrobial Resistance in Escherichia coli Isolates from Lettuce, Irrigation Water, and Soil 79. doi: 10.1128/AEM.01995-13.

Howard, Keya J.; Martin, Emily; Gentry, Terry; Feagley, Sam; Karthikeyan, Raghupathy (2016): Effects of Dairy Manure Management Practices on E. coli Concentration and Diversity. In Water, Air, & Soil Pollution 228 (1), p. 4. doi: 10.1007/s11270-016-3182-7.

Hubbard, L. E.; Givens, C. E.; Griffin, D. W.; Iwanowicz, L. R.; Meyer, M. T.; Kolpin, D. W. (2020): Poultry litter as potential source of pathogens and other contaminants in groundwater and surface water proximal to large-scale confined poultry feeding operations. In 0048-9697 735, p. 139459. doi: 10.1016/j.scitotenv.2020.139459.

Huijbers, P. M. C.; Graat, E. A. M.; van Hoek, Aham; Veenman, C.; Jong, M.C.M.de; van Duijkeren, E. (2016): Transmission dynamics of extended-spectrum β-lactamase and AmpC β-lactamase-producing Escherichia coli in a broiler flock without antibiotic use. In Preventive Veterinary Medicine 131, pp. 12–19. doi: 10.1016/j.prevetmed.2016.07.001.

Ingham, Steven; Losinski, Jill; Andrews, Matthew; Breuer, Jane; Breuer, Jeffry; Wood, Timothy; Wright, Thomas (2004): Escherichia coli Contamination of Vegetables Grown in Soils Fertilized with Noncomposted Bovine Manure: Garden-Scale Studies 70, pp. 6420– 6427. doi: 10.1128/AEM.70.11.6420-6427.2004.

Ingham, Steven C.; Fanslau Melody A.; Engel Rebecca A.; Breuer Jeffry R.; Breuer Jane E.; Wright Thomas H. et al.(2005): Evaluation of Fertilization-to-Planting and Fertilization-to-Harvest Intervals for Safe Use of Noncomposted Bovine Manure in Wisconsin Vegetable Production. In 0362-028X 68 (6), pp. 1134–1142. doi: 10.4315/0362-028X-68.6.1134.

Iooss, Bertrand; Da Veiga, Sebastien; Janon, Alexandre; Pujol, Gilles Iooss;, Maintainer Bertrand; Rcpp, LinkingTo et al. (2024): Package ‘sensitivity’. In R Package from https://cran.r-project.or/package=sensitivity.

Islam, Mahbub; Doyle, Michael; Phatak, Sharad; Millner, Pat; Jiang, Xiuping (2004): Persistence of Enterohemorrhagic Escherichia coli O157:H7 in Soil and on Leaf Lettuce and Parsley Grown in Fields Treated with Contaminated Manure Composts or Irrigation Water 67, pp. 1365–1370. doi: 10.4315/0362-028X-67.7.1365.

Iwu, Chidozie D.; Okoh Anthony I. (2019): Preharvest Transmission Routes of Fresh Produce Associated Bacterial Pathogens with Outbreak Potentials: A Review. In 1660-4601 16 (22). doi: 10.3390/ijerph16224407.

Jacobs, Kyle; Wind, Lauren; Krometis, Leigh-Anne; Hession, W. Cully; Pruden, Amy (2019): Fecal Indicator Bacteria and Antibiotic Resistance Genes in Storm Runoff from Dairy Manure and Compost-Amended Vegetable Plots. In 0047-2425 48 (4), pp. 1038–1046. doi: 10.2134/jeq2018.12.0441.

Jensen, Annette; Storm, C.; Forslund, Anita; Baggesen, Dorte; Dalsgaard, Anders (2013): Escherichia coli Contamination of Lettuce Grown in Soils Amended with Animal Slurry 76, pp. 1137–1144. doi: 10.4315/0362-028X.JFP-13-011.

Jensen, Dane A.; Friedrich Loretta M.; Harris Linda J.; Danyluk Michelle D.; Schaffner Donald W. (2015): Cross contamination of Escherichia coli O157:H7 between lettuce and wash water during home-scale washing. In Food Microbiology 46, pp. 428–433. doi: 10.1016/j.fm.2014.08.025.

Josephs-Spaulding, Jonathan; Krogh, Thøger Jensen; Rettig, Hannah Clara; Lyng, Mark; Chkonia, Mariam; Waschina, Silvio et al. (2021): Recurrent Urinary Tract Infections: Unraveling the Complicated Environment of Uncomplicated rUTIs. In Frontiers in Cellular and Infection Microbiology Volume 11 - 2021. Available online at https://www.frontiersin.org/journals/cellular-and-infection-microbiology/articles/10.3389/fcimb.2021.562525.

Kasparaviciene, Beatrice; Novoslavskij, Aleksandr; Aksomaitiene, Jurgita; Stankeviciene, Jurate; Kasetiene, Neringa; Sinkevicius, Romualdas; Malakauskas, Mindaugas (2025): Prevalence and Antimicrobial Resistance of ESBL E. coli in Early Broiler Production Stage and Farm Environment in Lithuania. In 2076-2607 13 (2). doi: 10.3390/microorganisms13020425.

Leistner, R.; Gürntke, S.; Sakellariou, C.; Denkel, L. A.; Bloch, A.; Gastmeier, P.; Schwab, F. (2014): Bloodstream infection due to extended-spectrum beta-lactamase (ESBL)-positive K. pneumoniae and E. coli: an analysis of the disease burden in a large cohort. In Infection 42 (6), pp. 991–997. doi: 10.1007/s15010-014-0670-9.

Literák, Ivan; Reitschmied, Tomas; Bujnáková, Dobroslava; Dolejska, Monika; Cizek, Alois; Bardon, Jan et al. (2012): Broilers as a Source of Quinolone-Resistant and Extraintestinal Pathogenic Escherichia coli in the Czech Republic. In Microbial drug resistance (Larchmont, N.Y.) 19. doi: 10.1089/mdr.2012.0124.

Lopez, K.; Getty, K.J.K.; Vahl, Christopher (2015): Validation of washing treatments to reduce Escherichia coli 0157:H7 and Salmonella spp. on the surface of green leaf lettuce and tomatoes 35, pp. 377–384.

Luyckx, Kaat; van Weyenberg, Stephanie; Dewulf, Jeroen; Herman, Lola; Zoons, J.; Vervaet, E. et al.(2015): On-farm comparisons of different cleaning protocols in broiler houses. In Poultry science 94. doi: 10.3382/ps/pev143.

Lyhs, Ulrike; Ikonen, Ilona; Pohjanvirta, Tarja; Raninen, Kaisa; Perko-Mäkelä, Päivikki; Pelkonen, Sinikka (2012): Extraintestinal pathogenic Escherichia coli in poultry meat products on the Finnish retail market. In Acta Veterinaria Scandinavica 54 (1), p. 64. doi: 10.1186/1751-0147-54-64.

MacKinnon, M. C.; Sargeant, J. M.; Pearl, D. L.; Reid-Smith, R. J.; Carson, C. A.; Parmley, E. J.; McEwen, S. A. (2020): Evaluation of the health and healthcare system burden due to antimicrobial-resistant Escherichia coli infections in humans: a systematic review and meta-analysis. In 2047-2994 9 (1), p. 200. doi: 10.1186/s13756-020-00863-x.

Mallioris, Panagiotis; Teunis, Gijs; Lagerweij, Giske; Joosten, Philip; Dewulf, Jeroen; Wagenaar Jaap A. et al.(2023): Biosecurity and antimicrobial use in broiler farms across nine European countries: toward identifying farm-specific options for reducing antimicrobial usage. In 0950-2688 151, e13. doi: 10.1017/S0950268822001960.

Mancini, John L. (1978): Numerical estimates of coliform mortality rates under various conditions. In Journal (Water Pollution Control Federation), pp. 2477–2484.

Mellata, Melha (2013): Human and Avian Extraintestinal Pathogenic Escherichia coli: Infections, Zoonotic Risks, and Antibiotic Resistance Trends. In Foodborne Pathogens and Disease 10. doi: 10.1089/fpd.2013.1533.

Mootian, Gabriel; Wu, Wen-Hsuan; Matthews Karl R. (2009): Transfer of Escherichia coli O157:H7 from soil, water, and manure contaminated with low numbers of the pathogen to lettuce plants 72 11, pp. 2308–2312. Available online at https://api.semanticscholar.org/CorpusID:21229382.

Moreno, Miguel A.; García-Soto, Silvia; Hernández, Marta; Bárcena, Carmen; Rodríguez-Lázaro, David; Ugarte-Ruíz, María; Domínguez, Lucas (2019): Day-old chicks are a source of antimicrobial resistant bacteria for laying hen farms. In Veterinary Microbiology 230, pp. 221–227. doi: 10.1016/j.vetmic.2019.02.007.

Nilsson, Oskar; Börjesson, Stefan; Landén, Annica; Bengtsson, Björn (2014): Vertical transmission of Escherichia coli carrying plasmid-mediated AmpC (pAmpC) through the broiler production pyramid. In J Antimicrob Chemother 69 (6), pp. 1497–1500. doi: 10.1093/jac/dku030.

O’Flaherty, E.; Borrego, C. M.; Balcázar, J. L.; Cummins, E. (2018): Human exposure assessment to antibiotic-resistant Escherichia coli through drinking water. In 0048-9697 616-617, pp. 1356–1364. doi: 10.1016/j.scitotenv.2017.10.180.

O’Flaherty, E.; Solimini, A.; Pantanella, F.; Cummins, E. (2019a): The potential human exposure to antibiotic resistant-Escherichia coli through recreational water. In 0048-9697 650 (Pt 1), pp. 786–795. doi: 10.1016/j.scitotenv.2018.09.018.

O’Flaherty, E.; Solimini, A. G.; Pantanella, F.; Giusti, M. de; Cummins, E. (2019b): Human exposure to antibiotic resistant-Escherichia coli through irrigated lettuce. In 0160-4120 122, pp. 270–280. doi: 10.1016/j.envint.2018.11.022.

Oikarainen, Paula E.; Pohjola Leena K.; Pietola Eeva S.; Heikinheimo, Annamari (2019): Direct vertical transmission of ESBL/pAmpC-producing Escherichia coli limited in poultry production pyramid. In Veterinary Microbiology 231, pp. 100–106. doi: 10.1016/j.vetmic.2019.03.001.

Okada, Elena; Young, Brian Jonathan; Pérez, Débora Jesabel; Pellegrini, María Celeste; Carciochi, Walter Daniel; Lavallén, Carla Mariela et al. (2024): Effect of on-farm poultry litter composting processes on physicochemical, biological, and toxicological parameters and reduction of antibiotics and antibiotic-resistant Escherichia coli. In Waste Management 174, pp. 310–319. doi: 10.1016/j.wasman.2023.12.005.

Ottoson, Jakob; Nyberg, Karin; Lindqvist, Roland; Albihn, Ann (2011): Quantitative Microbial Risk Assessment for Escherichia coli O157 on Lettuce, Based on Survival Data from Controlled Studies in a Climate Chamber. In Journal of food protection 74, pp. 2000– 2007. doi: 10.4315/0362-028X.JFP-10-563.

Pouillot, R.; Delignette-Muller, M. L.; Kelly, D. L.; Denis, J. B. (2016): The mc2d package. In R package 30.

Rawson, Thomas; Dawkins, Marian Stamp; Bonsall Michael B. (2019): A Mathematical Model of Campylobacter Dynamics Within a Broiler Flock. In 1664-302X Volume 10 - 2019. Available online at https://www.frontiersin.org/journals/microbiology/articles/10.3389/fmicb.2019.01940.

Robé, Caroline; Blasse, Anja; Merle, Roswitha; Friese, Anika; Roesler, Uwe; Guenther, Sebastian (2019): Low Dose Colonization of Broiler Chickens With ESBL-/AmpC-Producing Escherichia coli in a Seeder-Bird Model Independent of Antimicrobial Selection Pressure. In 1664-302X Volume 10 - 2019. Available online at https://www.frontiersin.org/journals/microbiology/articles/10.3389/fmicb.2019.02124.

Robé, Caroline; Daehre, Katrin; Merle, Roswitha; Friese, Anika; Guenther, Sebastian; Roesler, Uwe (2021): Impact of different management measures on the colonization of broiler chickens with ESBL- and pAmpC-producing Escherichia coli in an experimental seeder-bird model. In PLOS ONE 16 (1), e0245224. doi: 10.1371/journal.pone.0245224.

Robé, Caroline; Projahn, Michaela; Boll, Katrin; Blasse, Anja; Merle, Roswitha; Roesler, Uwe; Friese, Anika (2024): Survival of highly related ESBL- and pAmpC-producing Escherichia coli in broiler farms identified before and after cleaning and disinfection using cgMLST. In BMC Microbiology 24 (1), p. 143. doi: 10.1186/s12866-024-03292-7.

Sarnino, Nunzio; Basak, Subhasish; Collineau, Lucie; Merle, Roswitha (2025): Pathways of Escherichia coli transfer from animal manure: risks and mitigation in agriculture. In Frontiers in Public Health Volume 13 - 2025. Available online at https://www.frontiersin.org/journals/public-health/articles/10.3389/fpubh.2025.1568621.

Sharma Manan; Millner Patricia D.; Hashem Fawzy; Vinyard Bryan T.; East Cheryl L.; Handy Eric T. et al. (2019): Survival of Escherichia coli in Manure-Amended Soils Is Affected by Spatiotemporal, Agricultural, and Weather Factors in the Mid-Atlantic United States 85 (5), e02392.#x2010;18. doi: 10.1128/AEM.02392-18.

Siller, Paul; Daehre, Katrin; Thiel, Nadine; Nübel, Ulrich; Roesler, Uwe (2020): Impact of short-term storage on the quantity of extended-spectrum beta-lactamase–producing Escherichia coli in broiler litter under practical conditions. In 0032-5791 99 (4), pp. 2125– 2135. doi: 10.1016/j.psj.2019.11.043.

Solomon, Ethan; Yaron, Sima; Matthews, Karl: Transmission of Escherichia coli O157:H7 from Contaminated Manure and Irrigation Water to Lettuce Plant Tissue and Its Subsequent Internalization 68, pp. 397–400. doi: 10.1128/AEM.68.1.397-400.2002.

Song, Jihyun; Oh, Sung-Suck; Kim, Junghee; Shin, Jinwook (2020): Extended-spectrum β-lactamase-producing Escherichia coli isolated from raw vegetables in South Korea. In Scientific Reports 10 (1), p. 19721. doi: 10.1038/s41598-020-76890-w.

Sowah, Robert; Bradshaw, Kenneth; Snyder, Blake; Spidle, David; Molina, Marirosa (2020): Evaluation of the soil and water assessment tool (SWAT) for simulating E. coli concentrations at the watershed-scale 746, p. 140669. doi: 10.1016/j.scitotenv.2020.140669.

Stromberg, Zachary R.; Johnson James R.; Fairbrother John M.; Kilbourne, Jacquelyn; van Goor, Angelica; Curtiss, Roy, 3rd; Mellata, Melha (2017): Evaluation of Escherichia coli isolates from healthy chickens to determine their potential risk to poultry and human health. In PLOS ONE 12 (7), e0180599. doi: 10.1371/journal.pone.0180599.

Subirats, Jessica; Murray, Roger; Yin, Xiaole; Zhang, Tong; Topp, Edward (2021): Impact of chicken litter pre-application treatment on the abundance, field persistence, and transfer of antibiotic resistant bacteria and antibiotic resistance genes to vegetables. In 0048-9697 801, p. 149718. doi: 10.1016/j.scitotenv.2021.149718.

Suzuki, Yoshihiro; Horita, Tomoyuki; Nishimura, Emi; Xie, Hui; Tamai, Soichiro; Kobayashi, Ikuo et al. (2024): Crop contamination evaluation by antimicrobial-resistant bacteria via livestock waste compost-fertilized field soil. In 0304-3894 480, p. 135987. doi: 10.1016/j.jhazmat.2024.135987.

Thomas, C.; Idler, C.; Ammon, C.; Amon, T. (2024): Applied Research Note: Survival of Escherichia coli and temperature development during composting of chicken manure with a typically low carbon/nitrogen ratio and moisture content. In Journal of Applied Poultry Research 33 (2), p. 100402. doi: 10.1016/j.japr.2024.100402.

Tofani, Silvia; Albini, Elisa; Blasi, Francesca; Cucco, Lucilla; Lovito, Carmela; Maresca, Carmen et al. (2022): Assessing the Load, Virulence and Antibiotic-Resistant Traits of ESBL/Ampc E. coli from Broilers Raised on Conventional, Antibiotic-Free, and Organic Farms. In Antibiotics 11 (11). doi: 10.3390/antibiotics11111484.

Tsoularis, A.; Wallace, J. (2002): Analysis of logistic growth models. In Mathematical Biosciences 179 (1), pp. 21–55. doi: 10.1016/S0025-5564(02)00096-2.

Uhlig, Elisabeth; Olsson, Crister; He, Jiayi; Stark, Therese; Sadowska, Zuzanna; Molin, Goran et al. (2017): Effects of household washing on bacterial load and removal of Escherichia coli from lettuce and “ready‐to‐eat” salads. In Food Science & Nutrition 5. doi: 10.1002/fsn3.514.

van Overbeek, Leo; Duhamel, Marie; Aanstoot, Stefan; van der Plas, Carin Lombaers; Nijhuis, Els; Poleij, Leo et al. (2021): Transmission of Escherichia coli from Manure to Root Zones of Field-Grown Lettuce and Leek Plants. In 2076-2607 9 (11). doi: 10.3390/microorganisms9112289.

Weller, Daniel L.; Kovac, Jasna; Roof, Sherry; Kent David J.; Tokman Jeffrey I.; Kowalcyk, Barbara et al. (2017): Survival of Escherichia coli on Lettuce under Field Conditions Encountered in the Northeastern United States. In 0362-028X 80 (7), pp. 1214–1221. doi: 10.4315/0362-028X.JFP-16-419.

Wickham, Hadley; Averick, Mara; Bryan, Jennifer; Chang, Winston McGowan;, Lucy D’Agostino; François, Romain et al. (2019): Welcome to the Tidyverse. In Journal of open source software 4 (43), p. 1686.

Zhang, Yangjunna; Schmidt John W.; Arthur Terrance M.; Wheeler Tommy L.; Zhang, Qi; Wang, Bing (2022): A Farm-to-Fork Quantitative Microbial Exposure Assessment of β-Lactam-Resistant Escherichia coli among U.S. Beef Consumers. In 2076-2607 10 (3). doi: 10.3390/microorganisms10030661.

